# Energy Insecurity Influences Urban Outdoor Air Pollution Levels during COVID-19 Lockdown in South-Central Chile

**DOI:** 10.1101/2021.04.03.21254639

**Authors:** Aner Martinez-Soto, Constanza Avendaño-Vera, Alex Boso, Alvaro Hofflinger, Matthew Shupler

## Abstract

**Introduction:** In south-central Chile, outdoor air pollution primarily originates as household air pollution from wood burning for heating. The effect of COVID-19 lockdowns on ambient air pollution levels in urban south-central Chile may therefore be different from trends observed in cities where transportation and industrial emission sources dominate.

**Methods:** This quasi-experimental study compares hourly fine (PM_2.5_) and coarse (PM_10_) particulate matter measurements from six air monitoring stations (three reference grade beta attenuation monitors and three low-cost SPS30 sensors) in commercial and low or middle-income residential areas of Temuco and Padre Las Casas, Chile between March-September 2019 and 2020 (spanning COVID-19 lockdown).

**Results:** In Padre Las Casas, average outdoor PM_2.5_ concentrations peaked above 100 ug/m^3^ from 8-10 pm during winter (May-August) 2019 and 2020, when wood burning is common. During COVID-19 lockdown, average monthly ambient PM_2.5_ concentrations in a commercial and middle-income residential area of Temuco were up to 50% higher (12 µg/m^3^ to 18 µg/m^3^) and 59% higher (22 µg/m^3^ to 35 µg/m^3^) than 2019 levels, respectively. Conversely, PM_2.5_ levels decreased by up to 52% (43 µg/m^3^ to 21 µg/m^3^) in low-income neighborhoods. The night-time (8 pm-9 am) mass percent of PM_10_ that was PM_2.5_ during strict quarantine (April 2020) increased by 48% above April 2017-2019 proportions (50% to 74%) in a commercial area of Temuco.

**Conclusions:** Wood burning for home heating was responsible for a significantly higher proportion of ambient PM_2.5_ pollution in commercial areas and middle-income neighborhoods of Temuco during COVID-19 lockdown, compared to winter months in 2019. Constrastingly, energy insecure households likely refrained from wood heating during lockdown, leading to PM_2.5_ concentration declines. To reduce the double burden of ambient air pollution and energy insecurity in south-central Chile, affordability of clean heating fuels (e.g. electricity, liquefied petroleum gas) should be a policy priority.

## INTRODUCTION

Chile has undergone rapid economic growth over the last 30 years and was classified as a high-income country by the World Bank in 2013 (Pino et al., 2015). Yet, with significant levels of labor instability and informality (Ministerio de Desarrollo Social [MIDESO], 2018), income inequality in Chile is the highest among Organization for Economic Co-operation and Development (OECD) member countries (M Becerra et al., 2018). The highest poverty rates in Chile are in the south-central region, where many families live in subsidized, low quality housing with inadequate thermal insulation. Many of these households are energy insecure, being unable to afford thermally comfortable indoor conditions in winter months due to a combination of heating fuel prices, energy consumption needs and building properties (Pérez-Fargallo et al., 2020). In this region, households commonly burn wood in higher efficiency chimney stoves (leña) for cooking and heating (rural areas) (Shupler et al., 2020a) or only heating (urban areas) (R Reyes et al., 2015; René Reyes et al., 2019). Wood is the most popular energy source due to cultural preferences (Reeve et al., 2013) and affordability (wood is 4-5 times cheaper per unit of energy than cleaner sources (e.g. electricity or liquefied petroleum gas) (Martinez-Soto and Jentsch, 2019). The wood energy sector also plays a key role in livelihoods; over 60,000 people in south-central Chile are employed in supply chain and maintenance services related to woodstove heating (R Reyes et al., 2015).

In the cities of Temuco (capital of Araucanía) and Padre Las Casas in south-central Chile, approximately 80% of the 270,000 inhabitants rely on wood for heating (MMA, 2018). Poor quality housing construction in some residential areas of the cities allows household air pollution generated from wood burning to infiltrate the outdoor environment, significantly contributing to ambient air pollution (Chafe et al., 2015; Lai et al., 2019); this phenomen occurs in many low- and middle-income nations in Sub-Saharan Africa and Asia, where cooking with wood and other biomass is common (CDT, 2010; Huang et al., 2021). Like most cities in Chile, Temuco and Padre Las Casas are located in a valley between the Andes and coastal Cordillera mountains, leading to temperature inversion, limited rainfall, minimal air circulation and creating two of the highest polluted cities in North and South America (Martinez-Soto and Jentsch, 2019).

Burning firewood for heating has been shown to contribute 82% and 94% of total annual emissions of coarse (PM_10_) and fine (PM_2.5_) particulate matter in Temuco, respectively (Boso et al., 2018). As an estimated 93% of the PM from firewood burning is PM_2.5_ (Bravo-Linares et al., 2016), the average fine fraction of PM_10_ (mass proportion of PM_2.5_ in PM_10_) is roughly double in Temuco (80–90%) than Santiago (30–60%) (Sanhueza et al., 2009), the capital of Chile, where most ambient air pollution originates from traffic (Tsapakis et al., 2002). Because of the high levels of PM_2.5_ found in woodsmoke, ambient PM_2.5_ concentrations in Temuco usually exceed the World Health Organization average annual PM_2.5_ target level (25 μg/m^3^) during more than one-third of days in a typical year (120 days) (Díaz-Robles et al., 2008; Molina et al., 2017). In addition to PM_2.5_ being more harmful than PM_10_ for human health due to its ability to travel deeper into the lungs (Pope et al., 2015), the chemical composition of PM from woodsmoke places residents of Temuco at higher risk for respiratory and cardiovascular mortality, relative to individuals living in Chilean cities, where transporation and industry are the main air pollution sources (Díaz-Robles et al., 2014).

In urban areas with emissions primarily originating from transporation and industry, lockdowns enforced in 2020 to control spread of the Coronavirus Disease 2019 (COVID-19) (Hasnain et al., 2020; Thu et al., 2020) simultaneously reduced ambient air pollution levels (Collivignarelli et al., 2020; Gayen et al., 2021). For example, in Delhi, India, the average concentration of PM_10_ and PM_2.5_ decreased by 57% and 33%, respectively, compared to the previous three years (Mahato et al., 2020). In Zaragoza, Spain and Rome, Italy average PM_2.5_ concentrations decreased by 58% and 24%, respectively, during March 2020 compared to February of the same year (Chauhan and Singh, 2020).

In south-central Chile, where ambient air pollution originates primarily from residential wood burning, the effect of COVID-19 lockdown on urban air quality may be different. As COVID-19 containment measures may have altered wood heating patterns during winter months of 2020 due to more time spent at home and loss of employment or reductions in income, household and ambient air pollution levels may not have declined as in other cities (Querol et al., 2021). This study therefore assesses the effect of COVID-19 lockdown on ambient PM_2.5_ and PM_10_ concentrations in Temuco and Padre Las Casas from March-September 2020. As exposure to PM_2.5_ from wood burning is associated with several cardiovascular and respiratory diseases (Peña et al., 2017; Pope et al., 2015; Siddharthan et al., 2018), and increased ambient PM_2.5_ and PM_10_ levels have been associated with higher COVID-19 transmission (Coccia, 2020; Ogen, 2020; Zhu et al., 2020) and mortality (Setti et al., 2020; Wu et al., 2020), an understanding of the spatial variation in urban ambient air pollution levels in south-central Chile during COVID-19 lockdown can inform future policy measures aimed at protecting public health.

## METHODS

### COVID-19 Lockdown in Chile

On March 16, 2020, the Chilean government announced the closure of all shopping malls, daycares, schools, and universities. Two days later, borders were closed to international travel, and a national emergency was declared; a night-time curfew was instated on March 22, 2020 (Tariq et al., 2021). COVID-19 lockdown measures were categorized based on the number of infected persons in a localized area: (1) Quarantine (2) Transition (3) Preparation (4) Initial opening (Cuadrado et al., 2020). The strictest “quarantine” phase, in which movement was restricted and only essential businesses remained open, occurred from March 28 to April 29, 2020. A “transition” period followed from April 30 to July 23, 2020 in Temuco, which allowed unrestricted movement Monday-Friday for nonessential purposes. A “preparation” phase included free movement every day of the week and allowed gatherings of up to 25 people in closed spaces and up to 50 people in open spaces. In the “initial opening” period, which occurred from July 24 to October 2, 2020 in Temuco, face-to-face classes with up to 25 students in schools were additionally allowed.

### Air pollution measurements

A quasi-experimental study was conducted by comparing hourly PM_2.5_ and PM_10_ concentrations from March 1st to September 30th for 2019 and the same period in 2020 (during COVID-19 lockdown) using six air monitoring stations located throughout Temuco and Padre Last Casas. Three stations contained reference grade Met One beta attenuation monitors (BAM-1020, Met One Instruments, Oregon, USA) (Gobeli et al., 2008) maintained by the Chilean Ministry of the Environment (MMA), and three stations consisted of low-cost Sensirion SPS30 sensors (Bruce-Keller et al., 2018; Maag et al., 2018).

The limit of detection of the beta attenuation monitors is 4.0 µg/m^3^ and all measurements from the beta attenuation monitors used in this study are publicly available (http://sinca.mma.gob.cl/) via the National Air Quality Information System (SINCA) (SINCA, 2015). The SPS30 sensors are part of a larger network of 30 sensors throughout Temuco, developed by the Center for Software Engineering Studies at Universidad de La Frontera (UFRO) (“AIRE CEIUFRO,” 2019) with support from the municipality of Temuco, that provide real-time air quality data to residents via a mobile and web application called “AIRE Temuco” (http://aire.ceisufro.cl/#/dashboard).

Measurements from the three SPS30 sensors, the most accurate sensors in the “AIRE Temuco” network, were calibrated to a Grimm laser aerosol spectrometer 11-E (Grimm Technologies, Ainring, Germany) over an 8-hour period on December 24, 2019 (midway through the study period) in an airtight chamber. The efficiency of the laser aerosol spectrometer meets the European standards (EN12341 and EN14907) for PM_10_ and PM_2.5_, respectively. The coefficient of determination (R^2^) between the sensors and the laser aerosol spectrometer during the 8-hour calibration period was 0.92 for PM_2.5_ and 0.96 for PM_10_ (see Figures S3 and S4 in supplement for calibration curves). Field collocation of similar sensors and beta attenuation monitors in Santiago, Chile also showed relatively high correlation among 24-hour average PM_2.5_ concentrations (R^2^=0.63–0.87) (Tagle et al., 2020).

### Heating degree days

As colder winter temperatures generally lead to higher rates of wood burning for heating, a heating degree day (HDD) approach (Dickson, 1961; Liddell et al., 2016) was used to assess the impact of ambient temperature changes on PM_2.5_ and PM_10_ measurements between study years. HDD, a proxy of energy demand, compares the mean (average of the high and low) outdoor temperature recorded for a location to a standard temperature, in this case 15° Celsius (C) (based on calculations of thermal building performance from the International Organization for Standardization (ISO 15927-6) (British Standard Institution, 2008)). For example, HDD of 12 corresponds to an outdoor temperature of 3° C because it is 12 degrees below 15° C.

The HDD was calculated historically from 2009 to 2020, with average temperatures obtained from the Chilean Meteorological Directorate - Climate Services (“Dirección Meteorológica de Chile – Servicios Climáticos, Dirección General De Aeronáutica Civil (DGAC). (https://climatologia.meteochile.gob.cl/).,” n.d.). All temperatures used in the HDD calculation were converted to Kelvin, which is the standard (Dirección Meteorológica de Chile – Servicios Climáticos, Dirección General De Aeronáutica Civil (DGAC)); HDD are reported in units of Kelvin days (Kd) (equation 1).

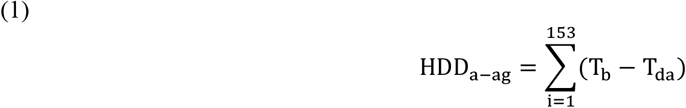

Where:

HDD_*a*−*ag*_: sum of heating degree days for April-August period [Kelvin days (Kd)].

T_b_ : base temperature [°C]. T_b_ =15°C

T_da_ : daily average temperature [°C]. T_da_ >15°C is not included in the HDD calculation.

### Statistical analysis

As the distribution of PM measurements was right skewed, all data were log transformed; geometric means (referred to as ‘mean’ from this point forward) are reported. Measurements from the SPS30 sensors, obtained every 10-minutes, were aggregated into hourly and monthly means. The fine fraction of PM_10_ (mass percent of PM_10_ that is PM_2.5_) was calculated by dividing the mean PM_2.5_ concentration recorded at a monitoring station by the mean PM_10_ concentration from the same station, over a given time period. Analysis and generation of figures were conducted in SPSS Statistics for Windows version 25.0 (IBM Corp, 2017) and R version 3.5.1 (R Core Team, 2017).

## RESULTS

The six monitors (Figure 1) are located in low-income residential (Pedro Valdivia, Pueblo Nuevo, Padre Las Casas), middle-income residential (Las Encinas) and commercial (Centro, Ñielol) areas (Table 1).

**Table 1.** Description of air monitor locations

| Monitor type | Location | Latitude | Longitude | Set up date | Residential or commercial area |
| --- | --- | --- | --- | --- | --- |
| Beta attenuation | Las Encinas | -38.71148 | -72.58042 | August 2012 | Middle-income residential |
|  | Padre Las Casas | -38.74927 | -72.59931 | June 2012 | Combination of low-income residential and commercial |
|  | Ñielol | -38.73326 | -72.62122 | May 2017 | Commercial |
| SPS30 sensor (collocated with beta attenuation monitors) | Pedro de Valdivia | -38.71076 | -72.62667 | April 2019 | Low-income residential |
|  | Centro | -38.73731 | -72.58939 | April 2019 | Commercial (city center) |
|  | Pueblo Nuevo | -38.71805 | -72.56433 | April 2019 | Low-income residential |

**Figure 1.**
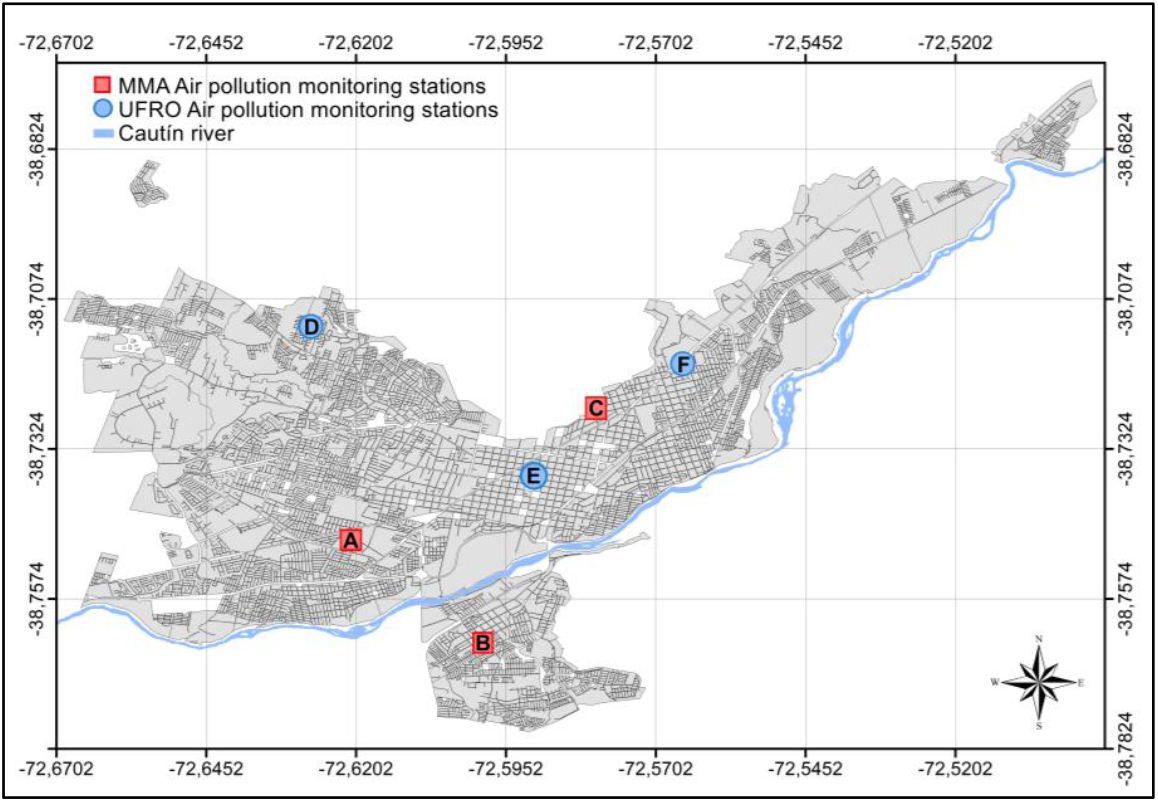
Air pollution monitoring stations in study setting: (A) Las Encinas, (B) Padre Las Casas, (C) Ñielol, (D) Pedro de Valdivia, (E) Centro, (F) Pueblo Nuevo.

### Relationship between heating degree days and ambient air pollution

At the two longest-running monitoring stations in Temuco (Las Encinas and Padre Las Casas) (Table 1), differences in number of heating degree days (HDD) were highly correlated (r=0.78-0.85) with average monthly PM_2.5_ concentrations during July from 2009-2020 (Las Encinas) and 2013-2020 (Padre Las Casas) (Figure 2).

**Figure 2.**
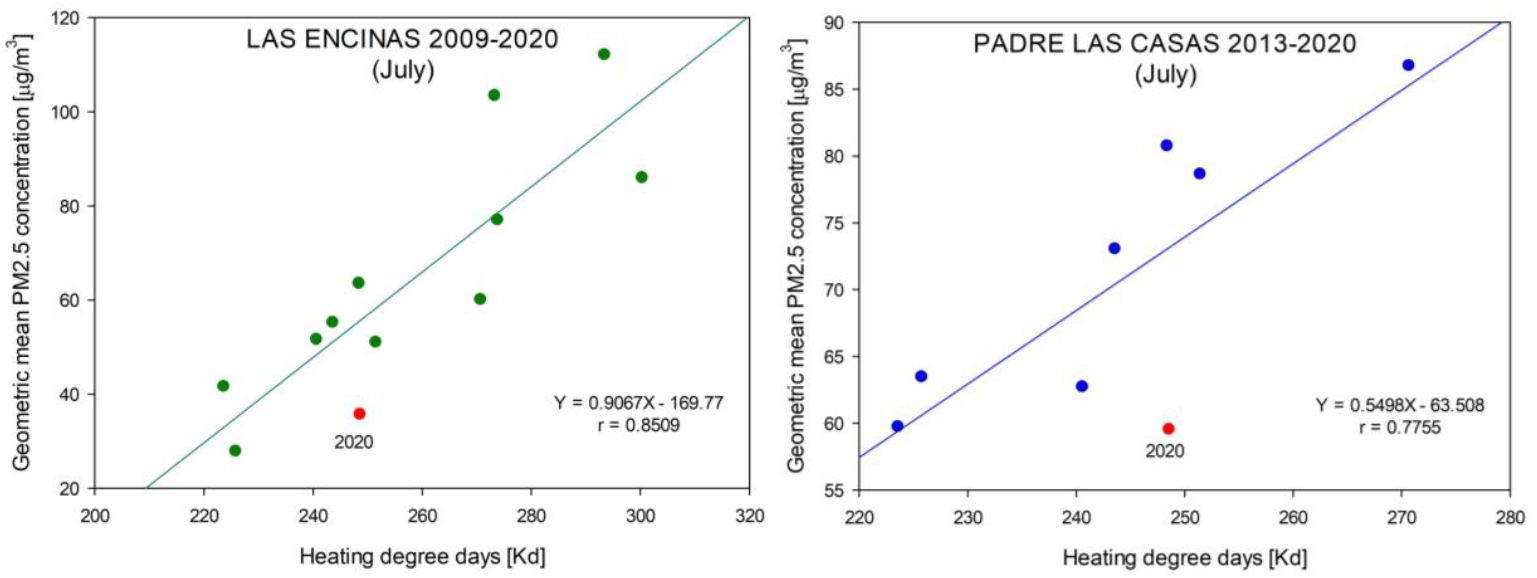
Average PM_2.5_ concentrations and heating degree days for July 2009-2020 (Las Encinas) and July 2013-2020 (Padre Las Casas)

The total number of HDD in April-August from 2009-2020 ranged from approximately 850 Kd (2015) to 1,100 Kd (2010), with an overall average of nearly 1000 Kd (Figure 3). As the number of HDD in 2019 and 2020 were similar (approximately 950 Kd; singaling slightly warmer winters than the overall average), air pollution measurements in this study were compared between 2019 and 2020 to minimize any confounding effect of winter temperature fluctuations on ambient PM concentrations.

**Figure 3.**
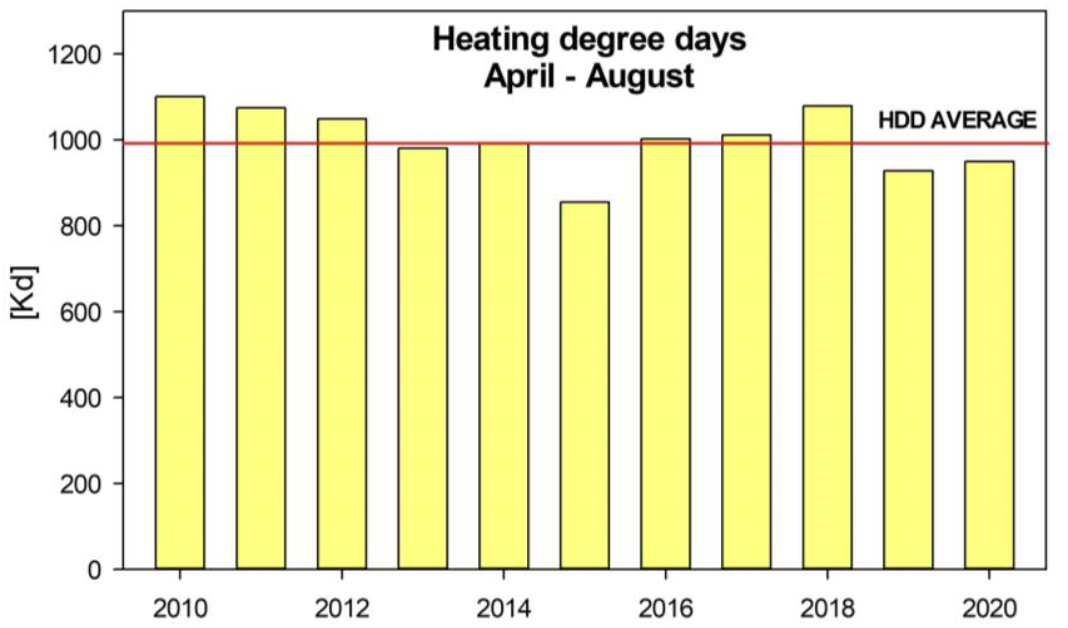
Number of heating degree days in Temuco, Chile during April-August 2009-2020

In 2019, average monthly PM_2.5_ concentrations during the winter months of June-August in low-income residential areas, including Padre Las Casas (36-47 µg/m^3^), Pedro Valdivia (40-42 µg/m^3^) and Pueblo Nuevo (42-53 µg/m^3^) were approximately twice as high as concentrations in commercial areas of Temuco (Nielol (12-15 µg/m^3^) and Centro (26-29 µg/m^3^)) (Table 2). Apart from the monitoring station in Nielol, average monthly PM_2.5_ concentrations recorded at all monitoring stations exceeded the Chilean standard of 20 µg/m^3^ during each of the three winter months in 2019.

**Table 2.** Average monthly PM_2.5_ and PM_10_ concentrations during winter months in 2019 and 2020

|  | Nielol (commercial) |  |  |  |  |  | Padre Las Casas (mixed commercial and low-income residential) |  |  |  |  |  | Las Encinas (middle-income residential) |  |  |  |  |  |
| --- | --- | --- | --- | --- | --- | --- | --- | --- | --- | --- | --- | --- | --- | --- | --- | --- | --- | --- |
|  | Average PM<sub>2.5</sub> concentration (µg/m<sup>3</sup>) |  |  | Average PM<sub>10</sub> concentration (µg/m<sup>3</sup>) |  |  | Average PM<sub>2.5</sub> concentration (µg/m<sup>3</sup>) |  |  | Average PM<sub>10</sub> concentration (µg/m<sup>3</sup>) |  |  | Average PM<sub>2.5</sub> concentration (µg/m<sup>3</sup>) |  |  | Average PM<sub>10</sub> concentration (µg/m<sup>3</sup>) |  |  |
| Month | 2019 | 2020 | % change | 2019 | 2020 | % change | 2019 | 2020 | % change | 2019 | 2020 | % change | 2019 | 2020 | % change | 2019 | 2020 | % change |
| March | 3 | 3 | 0% | 22 | 16 | -23% | 9 | 7 | -22% | 26 | 18 | -31% | 9 | 6 | -34% | 24 | 17 | -29% |
| **April**<br>(quarantine) | 8 | 11 | 38% | 24 | 17 | -29% | 21 | 22 | 5% | 34 | 31 | -9% | 18 | 17 | -5% | 29 | 24 | -17% |
| **May**<br>(transition) | 15 | 21 | 40% | 29 | 27 | -7% | 37 | 43 | 16% | 46 | 46 | 0% | 22 | 35 | 59% | 34 | 36 | 6% |
| **June**<br>(transition) | 14 | 17 | 21% | 25 | 22 | -12% | 36 | 35 | -3% | 43 | 36 | -16% | 23 | 25 | 9% | 30 | 25 | -17% |
| **July**<br>(transition) | 12 | 18 | 50% | 21 | 23 | 10% | 43 | 41 | -5% | 49 | 44 | -10% | 20 | 25 | 25% | 31 | 25 | -19% |
| **August**<br>(opening) | 15 | 14 | -4% | 27 | 19 | -30% | 47 | 40 | -15% | 55 | 46 | -16% | 23 | 25 | 9% | 38 | 33 | -13% |
| **Sept.**<br>(opening) | 11 | 13 | 18% | 19 | 23 | 21% | 27 | 32 | 19% | 34 | 41 | 21% | 16 | 24 | 50% | 26 | 33 | 27% |
| **Mean:**<br>**overall** | 11 | 14 | 27% | 24 | 21 | -13% | 31 | 31 | 0% | 41 | 37 | -10% | 19 | 22 | 16% | 30 | 28 | -7% |
| **Mean:**<br>**lockdown**<br>**months<sup>a</sup>** | 12 | 17 | 42% | 25 | 22 | -12% | 34 | 35 | 3% | 43 | 39 | -9% | 21 | 25 | 19% | 31 | 28 | -10% |
| **Mean:**<br>**opening**<br>**months<sup>b</sup>** | 13 | 14 | 8% | 23 | 21 | -9% | 37 | 36 | -3% | 45 | 44 | -2% | 20 | 25 | 25% | 32 | 33 | 3% |
|  | **Centro (commercial)** |  |  |  |  |  | **Pedro Valdivia (low-income residential)** |  |  |  |  |  | **Pueblo Nuevo (low-income residential)** |  |  |  |  |  |
|  | **Average PM<sub>2.5</sub>**<br>**concentration**<br>**(µg/m<sup>3</sup>)** |  |  | **Average PM<sub>10</sub>**<br>**concentration**<br>**(µg/m<sup>3</sup>)** |  |  | **Average PM<sub>2.5</sub>**<br>**concentration**<br>**(µg/m<sup>3</sup>)** |  |  | **Average PM<sub>10</sub>**<br>**concentration**<br>**(µg/m<sup>3</sup>)** |  |  | **Average PM<sub>2.5</sub>**<br>**concentration**<br>**(µg/m<sup>3</sup>)** |  |  | **Average PM<sub>10</sub>**<br>**concentration**<br>**(µg/m<sup>3</sup>)** |  |  |
| **Month** | **2019** | **2020** | **%**<br>**change** | **2019** | **2020** | **%**<br>**change** | **2019** | **2020** | **%**<br>**change** | **2019** | **2020** | **%**<br>**change** | **2019** | **2020** | **%**<br>**change** | **2019** | **2020** | **%**<br>**change** |
| **May**<br>(transition) | 29 | 37 | 28% | 62 | 65 | 4% | 43 | 21 | -52% | 71 | 59 | -16% | 46 | 37 | -20% | 71 | 64 | -9% |
| **June**<br>(transition) | 26 | 31 | 19% | 59 | 65 | 9% | 42 | 24 | -43% | 69 | 60 | -13% | 42 | 27 | -36% | 68 | 61 | -9% |
| **July**<br>(transition) | 25 | 32 | 28% | 59 | 65 | 10% | 41 | 29 | -29% | 69 | 64 | -6% | 48 | 29 | -40% | 69 | 63 | -9% |
| **August**<br>(initial<br>opening) | 29 | 28 | -3% | 60 | 61 | 2% | 40 | 29 | -27% | 67 | 63 | -6% | 53 | 28 | -48% | 71 | 61 | -14% |
| **Mean:**<br>**overall** | 27 | 32 | 19% | 60 | 64 | 7% | 42 | 26 | -39% | 69 | 62 | -10% | 47 | 30 | -37% | 70 | 62 | -11% |
| **Mean:**<br>**lockdown**<br>**months<sup>c</sup>** | 27 | 33 | 22% | 60 | 65 | 8% | 42 | 22 | -48% | 70 | 61 | -13% | 45 | 31 | -32% | 69 | 63 | -9% |

- a. Lockdown months: April (quarantine) and May-July (transition) - b. Opening months: August-September (initial opening) - c. Lockdown months: May-July (transition)

### Effect of COVID-19 lockdown on fine particulate matter levels

During COVID-19 lockdown in April-July 2020 (‘quarantine’ and ‘transition’ periods), PM_2.5_ levels in commercial areas (Centro, Nielol) and a middle-income neighborhood (Las Encinas) of Temuco were an average of 19-42% higher than corresponding months in 2019 (Table 2). In a single month, average PM_2.5_ concentrations during the ‘transition’ period were up to 50% (July 2020 in Nielol) and 59% (May 2020 in Las Encinas) higher than the same month in 2019 (Table 2). Conversely, during the first month of the ‘initial re-opening’ phase (August 2020), average PM_2.5_ concentrations in the two commercial areas were 3-4% below August 2019 levels.

In Padre Las Casas, the highest night-time PM_2.5_ levels before and during COVID-19 lockdown were observed and remained relatively unchanged (Table 2), with average outdoor concentrations exceeding 100 µg/m^3^ from 8-10 pm during May-August of 2019 and 2020 (Figure 5). In contrast, average PM_2.5_ concentrations during the ‘transition’ phase (May-July 2020) of COVID-19 lockdown were substantially lower than 2019 levels in the low-income neighborhoods of Pedro Valdivia (−48%) and Pueblo Nuevo (−32%). The relative decline in PM_2.5_ levels between 2019 and 2020 in these two low-income communities in Temuco was greater during morning (7-11 am) and nighttime (5 pm-12 am) hours, when air pollution from heating is typically highest (Figure 4). Similarly, in the residential area of Las Encinas, increases in PM_2.5_ concentrations during lockdown (above baseline 2019 levels) were most visible at night during home heating hours (Figure 5). Conversely, the increase in PM_2.5_ concentrations in 2020 relative to 2019 in commercial areas (Centro, Nielol) of Temuco was more evenly distributed across all hours of the day (Figures 4 and 5).

**Figure 4.**
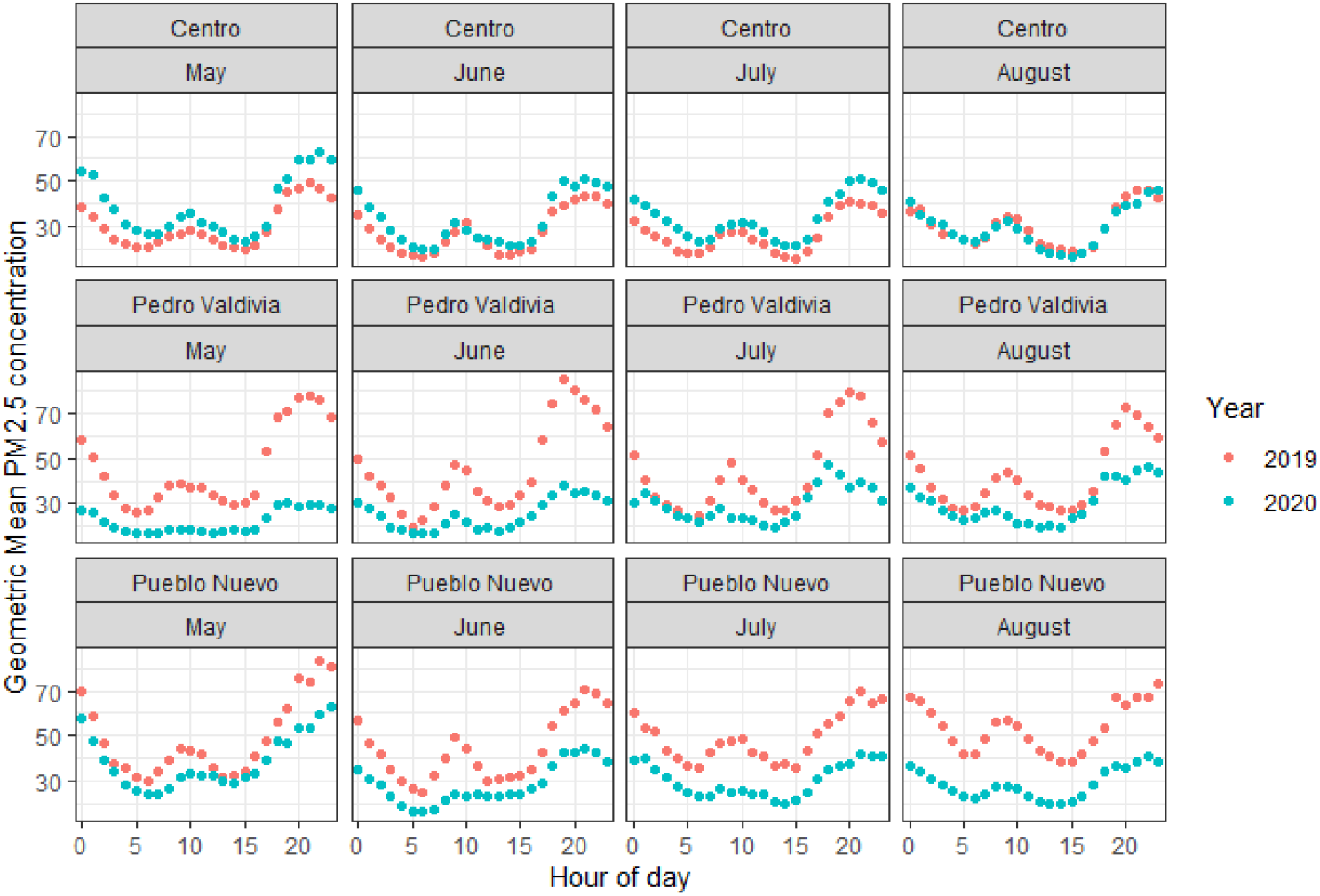
Hourly geometric mean PM_2.5_ concentrations obtained from SPS30 sensors during winter months of 2019 and 2020

**Figure 5.**
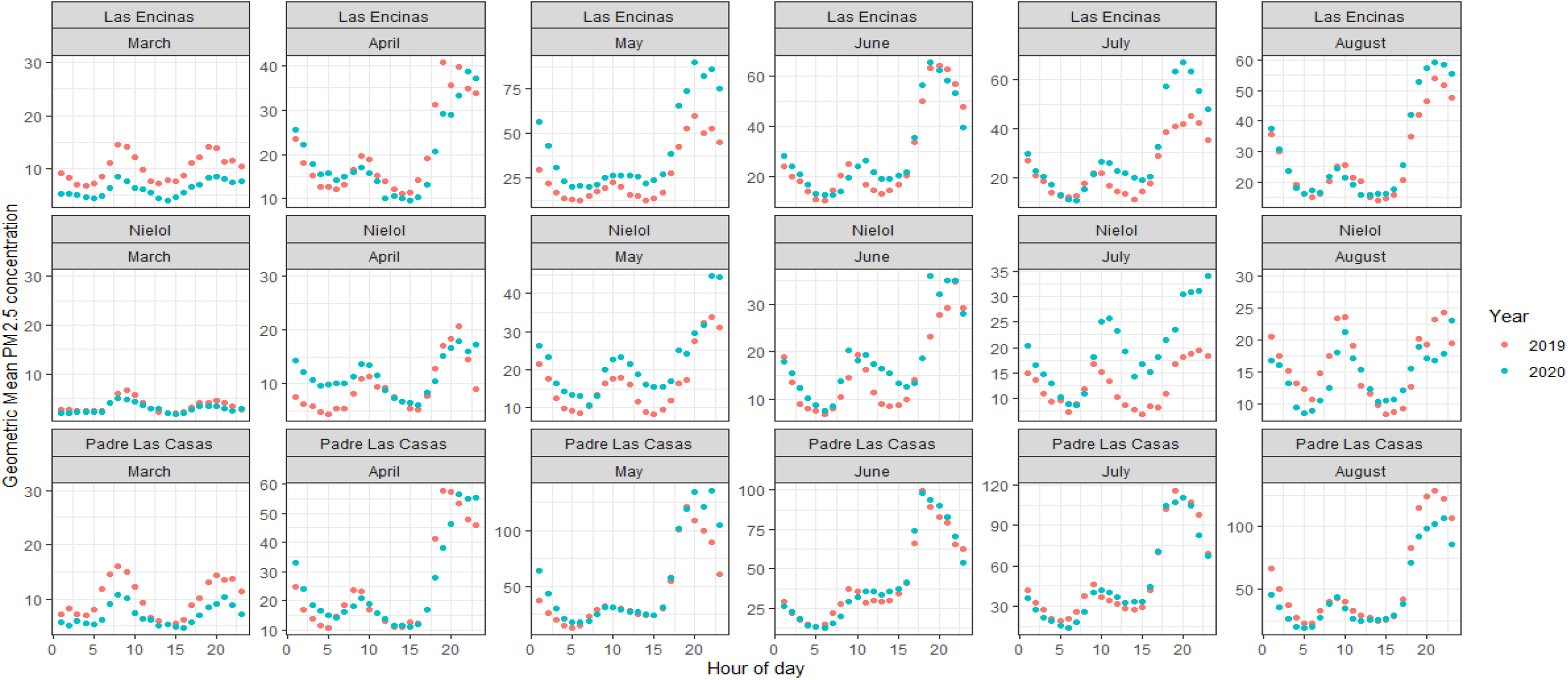
Hourly geometric mean PM_2.5_ concentrations obtained from beta attenuation monitors during winter months of 2019 and 2020

### Effect of COVID-19 lockdown on coarse particulate matter levels

Apart from the city center (Centro), average PM_10_ concentrations registered to all monitors were significantly lower during winter months of COVID-19 lockdown than in winter 2019 (Table 2; Figure 6). The largest relative decline in average monthly PM_10_ concentrations between 2019 and 2020 occurred in the commercial area of Nielol; the average PM_10_ concentration during the month of strict quarantine in Nielol (17 µg/m^3^) was 29% lower than April 2019 (24 µg/m^3^). A 7-12% lower average PM_10_ concentration in Nielol in 2020 compared to 2019 was maintained when restrictions were eased during the ‘transition’ phase in May and June (Table 2). The decline in PM_10_ concentrations in April 2020 relative to April 2019 in Nielol was visible at all hours of the day (Figure 7). In contrast, in the residential areas of Pedro Valdivia and Pueblo Nuevo, lower PM_10_ levels in 2020 compared with 2019 were most apparent during typical wood heating hours at night and early morning (Figures 6).

**Figure 6.**
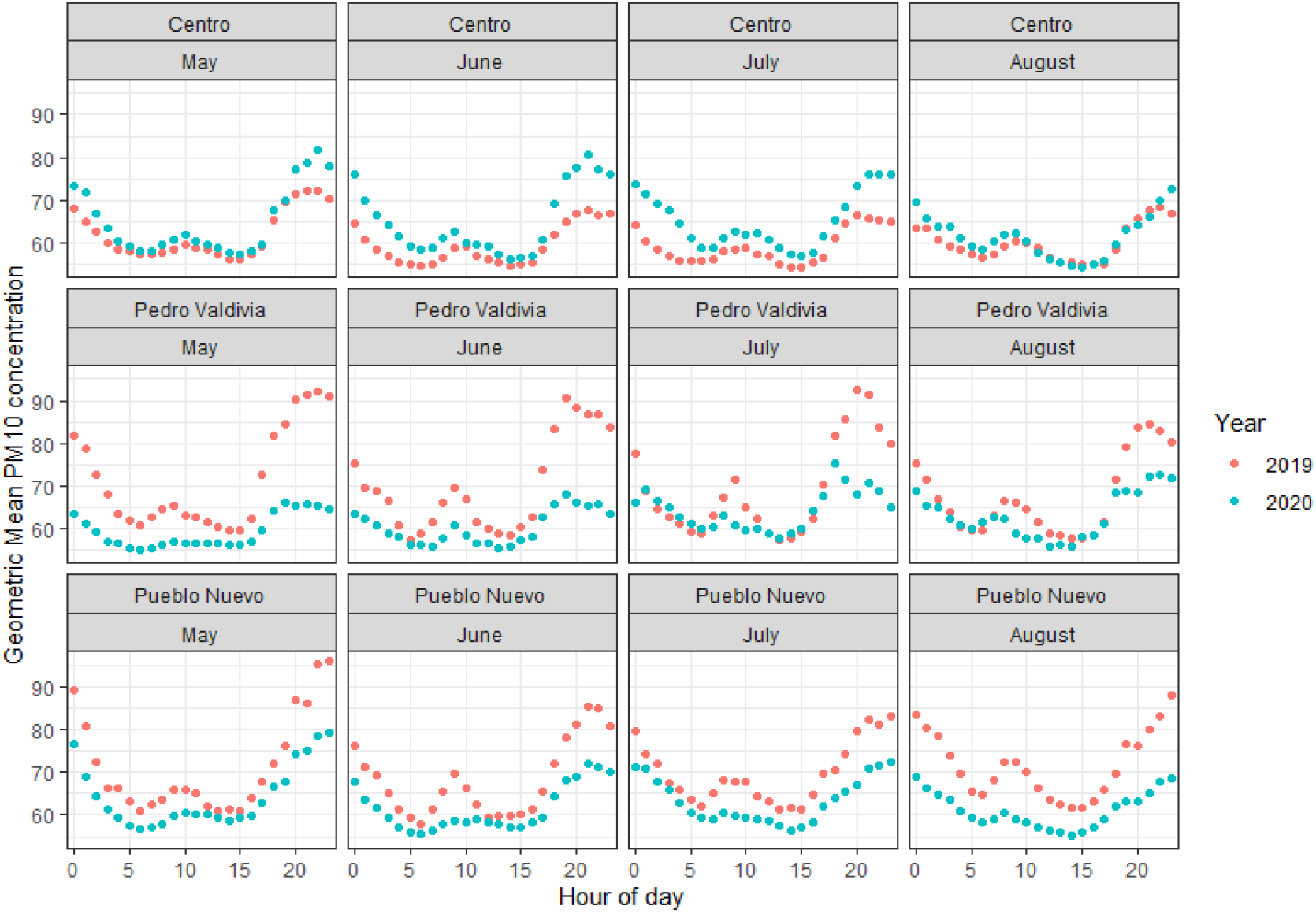
Hourly geometric mean PM_10_ concentrations obtained from SPS sensors during winter months of 2019 and 2020

**Figure 7.**
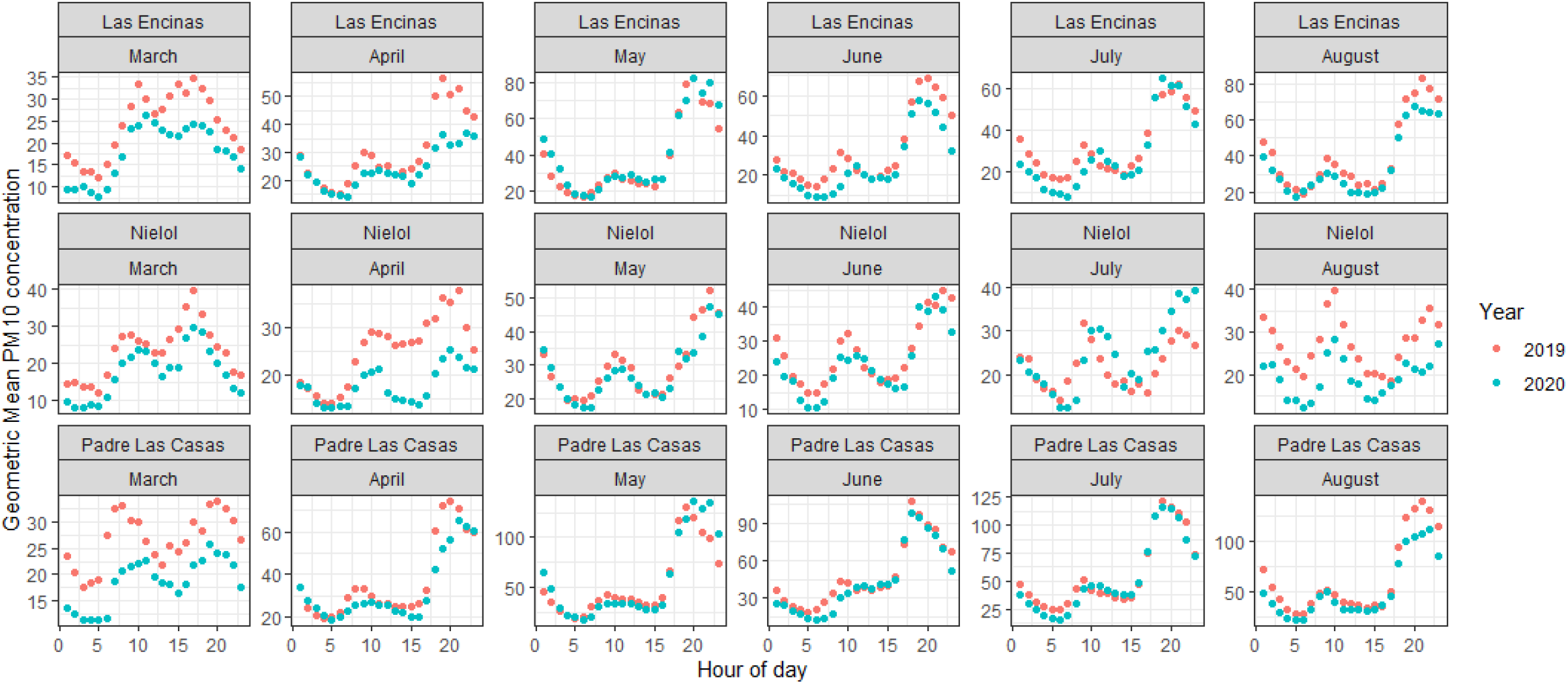
Hourly geometric mean PM_10_ concentrations obtained from beta attenuation monitors during winter months of 2019 and 2020

Because the size of particulate matter produced from residential wood burning is mostly PM_2.5_, monthly average PM_10_ concentrations decreased by a lesser extent (no more than 16%) in primarily residential neighborhoods compared with Nielol (Table 2).

### Effect of COVID-19 lockdown on fine fraction of PM_10_

In a commercial (Nielol) and middle-income residential area (Las Encinas), large increases in the fine fraction of PM_10_ occurred during the month of strict quarantine in April 2020 relative to April 2017-2019 (Figure 9); the fine fraction of PM_10_ obtained at these two monitoring stations at night in April 2020 was more than 40% higher than in April 2017-2019 (Table 3). During the daytime in April 2020, the increase in the fine fraction of PM_10_ above April 20187-2019 levels (32%) in Nielol was double that of Las Encinas (16%). During the transition period from May-July 2020, the fine fraction of PM_10_ during night hours in Las Encinas reached 100%, which was 33-40% greater than the recorded proportion in 2017-2019. In Nielol, the relative increase between the 2017-2019 and 2020 fine fraction of PM_10_ was conversely reduced by over 75% (48% to 11%) between the quarantine phase (April 2020) and the first month of the transition phase (May 2020) (Table 3). The percent increase in the fine fraction of PM_10_ in Las Encinas between 2020 and 2017-2019 levels was halved (35% to 17%) during the first month initial re-opening of Temuco in August 2020.

**Table 3.** Average and standard deviation (SD) of fine fraction of PM_10_ (%) registered to beta attenuation monitors during day and night hours during winter months (March-September) of 2017-2019 and 2020

| COVID-19 period | Month | Nielol (commercial) |  |  |  |  |  | Padre Las Casas (low-income residential) |  |  |  |  |  | Las Encinas (middle-income residential) |  |  |  |  |  |
| --- | --- | --- | --- | --- | --- | --- | --- | --- | --- | --- | --- | --- | --- | --- | --- | --- | --- | --- | --- |
|  |  | Day (9 am-8 pm) |  |  | Night (8 pm-9 am) |  |  | Day (9 am-8 pm) |  |  | Night (8 pm-9 am) |  |  | Day (9 am-8 pm) |  |  | Night (8 pm-9 am) |  |  |
|  |  | 2017-2019 avg | 2020 | % increase | 2017-2019 avg | 2020 | % increase | 2017-2019 avg | 2020 | % increase | 2017-2019 avg | 2020 | % increase | 2017-2019 avg | 2020 | % increase | 2017-2019 avg | 2020 | % increase |
| Pre-lockdown | Mar. | 19 (10) | 14 (4) | -26% | 27 (9) | 23 (4) | -15% | 59 (2) | 56 (5) | 0% | 67 (1) | 67 (4) | 0% | 23 (9) | 24 (6) | 4% | 40 (10) | 48 (5) | 20% |
| Quarantine | April | 41 (14) | 54 (9) | 32% | 50 (13) | 74 (5) | 48% | 78 (2) | 78 (6) | 0% | 84 (2) | 90 (5) | 7% | 48 (15) | 56 (12) | 16% | 65 (11) | 93 (6) | 43% |
| Transition | May | 69 (16) | 77 (4) | 12% | 70 (12) | 78 (11) | 11% | 88 (1) | 95 (3) | 8% | 88 (2) | 98 (3) | 11% | 60 (14) | 91 (5) | 57% | 70 (9) | 99 (2) | 41% |
|  | June | 72 (14) | 78 (5) | 8% | 71 (13) | 78 (7) | 10% | 92 (1) | 97 (2) | 5% | 91 (2) | 98 (3) | 8% | 72 (8) | 100 (1) | 39% | 74 (12) | 100 (0) | 33% |
|  | July | 68 (14) | 80 (4) | 18% | 72 (13) | 78 (8) | 8% | 91 (1) | 95 (2) | 4% | 92 (1) | 97 (2) | 5% | 68 (9) | 94 (6) | 38% | 75 (9) | 100 (0) | 35% |
| Initial opening | Aug. | 66 (11) | 75 (6) | 14% | 70 (9) | 75 (7) | 7% | 87 (1) | 91 (3) | 5% | 89 (2) | 95 (3) | 7% | 63 (11) | 76 (3) | 23% | 71 (11) | 83 (7) | 17% |
|  | Sept. | 50 (9) | 55 (9) | 10% | 64 (5) | 62 (3) | -3% | 79 (2) | 85 (4) | 8% | 87 (1) | 91 (3) | 5% | 51 (11) | 67 (5) | 31% | 70 (6) | 76 (4) | 9% |

In low-income neighborhoods of Pedro Valdivia and Pueblo Nuevo, the decrease in PM_2.5_ concentrations in 2020 compared with 2019 was much greater than the corresponding decrease in PM_10_ levels, leading to substantial reductions (30% or more) in the fine fraction of PM in 2020 (Figure 8). In Centro, a higher fine fraction of PM_10_ existed in 2020 compared with 2019, with a larger relative increase between the two years occurring during night and early morning hours (8 pm-9am) of the transition period (May-July) (Figure 8). During the first month of re-opening in August 2020, the average day and night fine fraction of PM_10_ in Centro switched to being below that of August 2019 (Table 4).

**Table 4.**
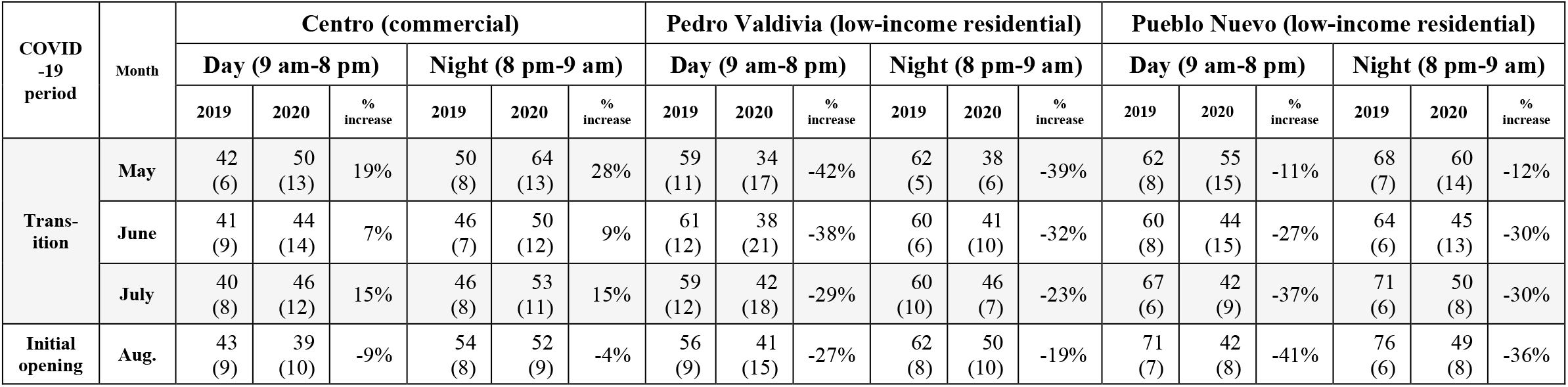
Average and standard deviation (SD) of fine fraction of PM_10_ (%) registered to SPS30 sensors during day and night hours during winter months (May-August) of 2019 and 2020

| COVID-19 period | Month | Centro (commercial) |  |  |  |  |  | Pedro Valdivia (low-income residential) |  |  |  |  |  | Pueblo Nuevo (low-income residential) |  |  |  |  |  |
| --- | --- | --- | --- | --- | --- | --- | --- | --- | --- | --- | --- | --- | --- | --- | --- | --- | --- | --- | --- |
|  |  | Day (9 am-8 pm) |  |  | Night (8 pm-9 am) |  |  | Day (9 am-8 pm) |  |  | Night (8 pm-9 am) |  |  | Day (9 am-8 pm) |  |  | Night (8 pm-9 am) |  |  |
|  |  | 2019 | 2020 | % increase | 2019 | 2020 | % increase | 2019 | 2020 | % increase | 2019 | 2020 | % increase | 2019 | 2020 | % increase | 2019 | 2020 | % increase |
| Transition | May | 42<br>(6) | 50<br>(13) | 19% | 50<br>(8) | 64<br>(13) | 28% | 59<br>(11) | 34<br>(17) | -42% | 62<br>(5) | 38<br>(6) | -39% | 62<br>(8) | 55<br>(15) | -11% | 68<br>(7) | 60<br>(14) | -12% |
|  | June | 41<br>(9) | 44<br>(14) | 7% | 46<br>(7) | 50<br>(12) | 9% | 61<br>(12) | 38<br>(21) | -38% | 60<br>(6) | 41<br>(10) | -32% | 60<br>(8) | 44<br>(15) | -27% | 64<br>(6) | 45<br>(13) | -30% |
|  | July | 40<br>(8) | 46<br>(12) | 15% | 46<br>(8) | 53<br>(11) | 15% | 59<br>(12) | 42<br>(18) | -29% | 60<br>(10) | 46<br>(7) | -23% | 67<br>(6) | 42<br>(9) | -37% | 71<br>(6) | 50<br>(8) | -30% |
| Initial opening | Aug. | 43<br>(9) | 39<br>(10) | -9% | 54<br>(8) | 52<br>(9) | -4% | 56<br>(9) | 41<br>(15) | -27% | 62<br>(8) | 50<br>(10) | -19% | 71<br>(7) | 42<br>(8) | -41% | 76<br>(6) | 49<br>(8) | -36% |

**Figure 8.**
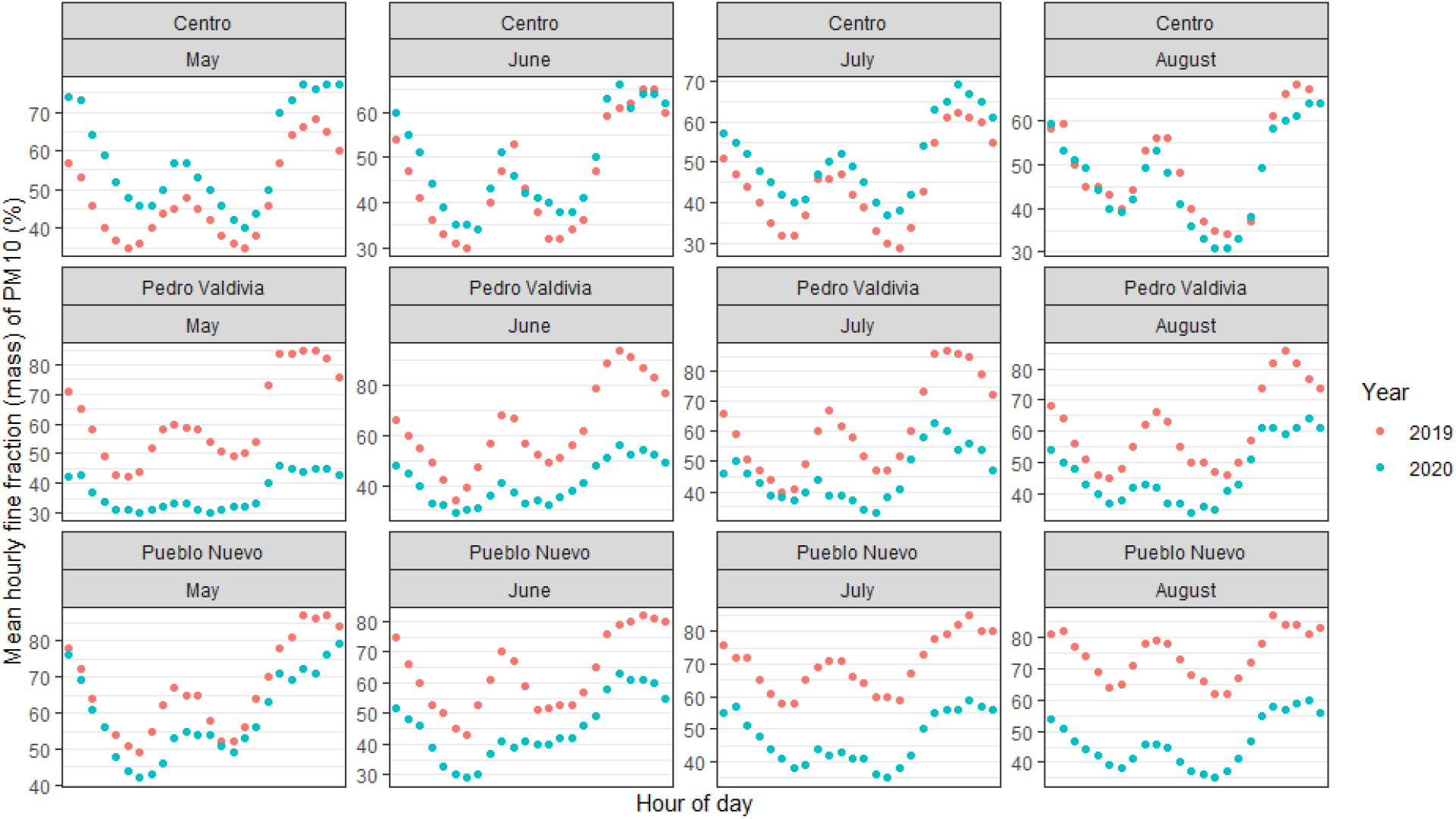
Mean hourly fine fraction (mass percent of PM_10_ that is PM_2.5_) in 2019 and 2020 obtained from SPS30 sensors

**Figure 9.**
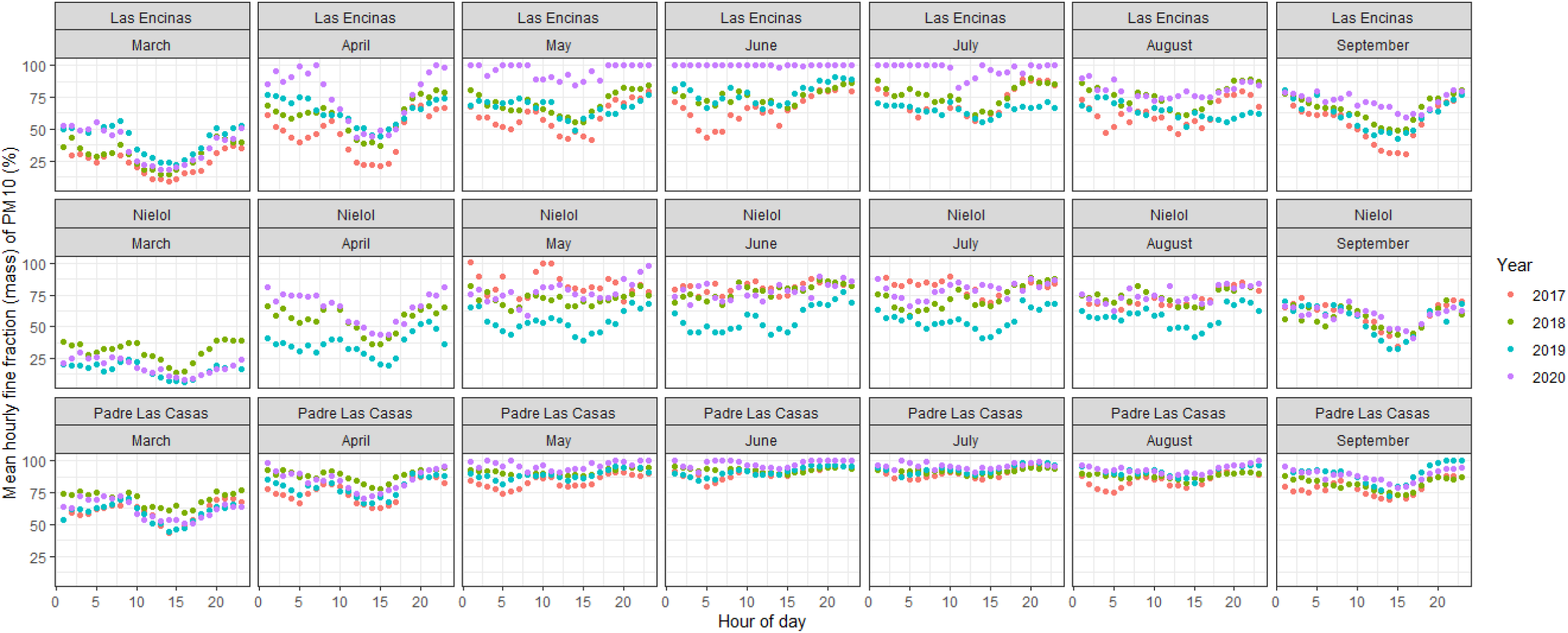
Mean hourly fine fraction (mass percent of PM_10_ that is PM_2.5_) in 2019 and 2020 obtained from beta attenuation monitors

## DISCUSSION

This quasi-experimental study presents a dichotomy of elevated ambient PM_2.5_ levels in commercial and middle-income neighborhoods and a decline in low-income areas of Temuco. With over 90% of PM emissions from residential wood burning consisting of PM_2.5_ (Díaz-Robles et al., 2014), the increase in average monthly outdoor PM_2.5_ concentrations in commercial areas by up to 50% yet decline in average monthly outdoor PM_10_ concentrations by up to 29% between 2019 and 2020 in Temuco was evidently driven by an increase in household wood burning during COVID-19 lockdown. A rise in average ambient PM_2.5_ levels in commercial areas in the study setting during COVID-19 lockdown is an anomalous result when compared with trends observed in other high-income countries, where urban ambient PM_2.5_ concentrations typically plummeted (by 24-159%) due to lower industrial emissions (Chauhan and Singh, 2020; Mahato et al., 2020).

### Health impacts of elevated ambient fine particulate matter levels

Higher PM_2.5_ concentrations during COVID-19 lockdown in several regions of Temuco is highly problematic from a health perspective, especially with possible increased length of exposure to household air pollution while confined at home (Chen et al., 2021). Research conducted in low and middle-income countries (LMICs) where there is high prevalence of cooking with wood and other polluting fuels demonstrates that elevated PM_2.5_ levels found in household air pollution places the population at increased risk for infectious, cardiovascular and respiratory diseases and mortality (Apte et al., 2015). As Chile had some of the highest rates of COVID-19 cases globally during April-August 2020 (Undurraga et al., 2021), it is possible that increased PM_2.5_ levels in Temuco may have exacerbated COVID-19 transmission (Fattorini and Regoli, 2020; Zhu et al., 2020). Older individuals (≥65 years), who have the greatest health risk from exposure to wood smoke (Díaz-Robles et al., 2015), and are more likely to suffer from severe symptoms of COVID-19, can be particularly vulnerable (Undurraga et al., 2021).

While most of the 2.3 million deaths due to household air pollution exposure in 2019 occurred among three billion individuals living in LMICs that cook with wood and other polluting fuels (Murray et al., 2020;  State of Global Air, 2020), this study suggests that household air pollution may have posed an increased health burden during COVID-19 lockdown in certain high-income countries where wood burning is prevalent. Recent studies conducted in Spain (Querol et al., 2021) and across Europe (Evangeliou et al., 2020) found that residential wood combustion was likely a more significant source of ambient emissions of PM_2.5_ and black carbon, a component of PM_2.5_ generated from wood combustion (Bond et al., 2013; Shupler et al., 2020a), during COVID-19 lockdown compared with years prior to the pandemic.

### Efforts to reduce ambient air pollution in Temuco

An existing Atmospheric Decontamination Plan (ADP) that aimed to reduce ambient PM_2.5_ concentrations in Temuco and Padre Las Casas was implemented by the local government in 2015. The ADP enforces cessation of residential wood-burning during specific ‘critical episodes’ (average 24-hour PM_2.5_ concentrations >80 μg/m^3^) (Table S1), with financial penalties levied for unabiding households (Boso et al., 2018). The ADP further subsidizes the thermal conditioning of homes and the replacement of older wood stoves with higher efficiency wood heaters, pellet stoves or kerosene stoves; nearly 8,000 wood stoves were replaced between 2011-2017 and 8,500 households were retrofitted between 2015-2017 under the program (Reyes et al., 2019). Although temporary bans on residential wood-burning during ‘critical episode’ periods has reduced short-term PM_2.5_ and PM_10_ concentrations by up to 17.5% and 9.2%, respectively (Mardones and Cornejo, 2020), the ADP has had limited effectiveness in reducing long-term PM_2.5_ concentrations (Schueftan and González, 2015). Replacement of all existing wood stoves with more efficient burning stoves may therefore not significantly achieve meaningful ambient PM_2.5_ concentration reductions in Temuco, especially with rising per capita energy consumption expected due to increasing household income (Jorquera et al., 2018).

The marked increase in PM_2.5_ levels during COVID-19 lockdown in Temuco underscores an imperative need for more robust policies that minimize wood burning for heating in the city. Providing households with affordable access to cleaner forms of energy (e.g. liquefied petroleum gas or electricity) for cooking and heating, which emit significantly lower PM_2.5_ emissions (Shupler et al., 2020a), could be a more effective policy component of ADPs (Bravo-Linares et al., 2016). Increased use of clean fuels would not only improve health outcomes from reductions in household and ambient PM_2.5_ concentrations, but also reduce localized rates of deforestation (Reyes et al., 2015; Schueftan and González, 2013) and potentially free up time for individuals to shift to other income-generating activities, due to lower time spent on stove maintence, chimney cleaning and storing, chopping and drying wood (Boso, A., Hofflinger, A., Garrido, J., & Álvarez, 2020).

### Socioeconomic heterogeneity in ambient particulate matter concentrations

The city of Padre las Casas has a larger indigenous population and higher poverty rates than Temuco (Barton and Ramírez, 2019). Pedro Valdivia, located on the outskirts of Temuco, and Pueblo Nuevo both have a high proportion of social housing, with families living in small residential condominiums and apartment buildings typically not larger than 30 square meters (Garín Contreras et al., 2009; Vergara, 2019). In these three residential areas, average PM_2.5_ concentrations decreased by an average of 32% (Pueblo Nuevo) and 48% (Pedro Valdivia) or did not increase (Padre las Casas) during COVID-19 lockdown despite more time confined at home and similar winter temperatures as 2019 (Figure 3). Thus, families in these three low-income neighborhoods likely used wood heating at a lower-than-usual rate due to financial strain (Reyes et al., 2015), particularly with over half (53%) of Chileans indicating household income declines or financial debt (60%) due to the COVID-19 pandemic, with a third (33%) reporting unemployment (Umaña-Hermosilla et al., 2020).

While household-level energy consumption patterns were not examined in this study, high rates of energy insecurity have been identified in Temuco prior to the pandemic (M Becerra et al., 2018). Over two-thirds (68%) of low-income households in southern Chile are estimated to keep their living room temperatures below the thermal comfort standard of 21°C established by WHO (Boso et al., 2018) during the winter months (R Reyes et al., 2019), accepting higher levels of thermal discomfort to reduce costs (Miguel Becerra et al., 2018; Boso, A., Hofflinger, A., Garrido, J., & Álvarez, 2020). Further, a similar decline in household energy consumption was found in an urban informal settlement in Nairobi, Kenya during COVID-19 lockdown, with many households cooking less frequently or switching from paid cooking fuels (e.g. liquefied petroleum gas) to gathering wood for free to offset significant income loss during lockdown (Shupler et al., 2020b). While socioeconomic and demographic differences exist between countries in Sub-Saharan Africa and Chile, both areas have a high population proportion working in the informal sector, with families in Temuco typically spending over 10% of household income on energy in the winter (Reyes et al., 2019) and those living in informal settlements in Nairobi spending up to 20% of monthly income on cooking fuels (Ahmed et al., 2015).

Additionally, as human mobility in Chile was reduced during COVID-19 lockdown to a lesser extent in lower socioeconomic municipalities (20-40% reduction) compared with more affluent areas (60-80% reduction) (Gozzi et al., 2020; Mena et al., 2021), the inability of families working in the informal sector to quarantine due to the need for daily wages may have partially contributed to lower home heating demands in low-income neighborhoods and therefore driven down PM_2.5_ levels (Jay et al., 2020).

These findings emphasize the negative downstream effects of the COVID-19 lockdown that disproportionately affected energy insecure households in Temuco, who likely did not have the income to pay for wood heating and additionally may have decided to continue their employment to support their families. Energy insecure households that refrained from heating their homes may have become more vulnerable to respiratory and cardiovascular stress due to living in unsafe, cold indoor temperatures for prolonged periods (Howden-Chapman and Chapman, 2012; Schueftan et al., 2016).

### Strengths and Limitations

This study leveraged publicly available data from low-cost sensors and regulatory-grade beta attenuation monitors strategically placed throughout commercial and low- and middle-income residential neighborhoods of Temuco and Padre Las Casas. Combining the monitoring networks enabled an examination of the differential impacts of COVID-19 lockdown on urban ambient air quality across neighborhoods with potentially distinct changes in energy consumption patterns, which may be attributed to variation in neighborhood-level socioeconomic status. While the accuracy of the low-cost sensors used in this study can be affected by meterological factors such as temperature and humidity, the sensors showed high agreement with the reference standard during calibration (Figures S3 and S4). Further, measurements were aggregated at monthly levels, thereby lowering the risk of measurement error impacting study findings (Malings et al., 2020). A heating degree day analysis revealed that PM_2.5_ concentration fluctuations during COVID-19 lockdown in 2020 relative to 2019 were likely not attributed solely to colder winter temperatures (Figure 2).

Household-level energy consumption patterns in Temuco and Padre Las Casas during COVID-19 lockdown were not assessed. However, given existing documented negative impacts of lockdowns on energy security (Shupler et al., 2020b) and the history of energy insecurity and inadequate maintenance of thermal comfort in the region prior to the pandemic (M Becerra et al., 2018; Boso et al., 2018), energy security was likely an influential factor in the dichotomy of ambient air pollution trends observed in Temuco during the study period. Data on household energy consumption behaviors in Temuco during lockdown can uncover policies needed to ensure low-income families are thermally protected during cold Chilean winters.

The spatial variation in the PM_2.5_ concentration patterns pre- and post-pandemic highlights the utility of having a dense sensor network across numerous residential and commercial districts in Temuco and Padre Las Casas, which can help identify priority areas for intervention where energy insecurity rates may be highest (e.g. Fuel Poverty Potential Risk Index) (Pérez-Fargallo et al., 2018). Promoting use of the existing 30-monitor network in Temuco as a public health tool, and potentially expanding the network in an economically viable manner using low-cost sensors, can empower residents to make informed decisions regarding the scheduling of outdoor activities using neighborhood-level, real-time air quality data (Kumar et al., 2015; Snyder et al., 2013).

## CONCLUSION

Differences in the main sources of ambient air pollution emissions between Temuco and Padre Las Casas and other urban areas in high-income countries led to vastly diverse impacts on urban air quality due to COVID-19 lockdown. Similar to findings in certain regions of Europe (Evangeliou et al., 2020; Querol et al., 2021), higher average outdoor PM_2.5_ concentrations in commercial and more affluent residential areas of Temuco during COVID-19 lockdown relative to 2019 due to higher rates of wood burning while confined indoors may have increased the risk of adverse cardiovascular and respiratory outcomes. Conversely, substantial reductions in ambient PM_2.5_ levels in low-income neighborhoods between 2019 and 2020 suggests lower-than-usual maintainenance of thermal comfort during lockdown, which can similarly lead to higher risk of adverse cardiovascular and respiratory events among energy insecure families. These results underscore the need for clean sources of household energy (e.g. electricity, liquefied petroleum gas) to be made affordable and accessible (e.g. via clean fuel subsidies and improved energy infrastructure) to avoid a double burden of both unsafe ambient air pollution levels and cold indoor residential temperatures during winter. More aggressive governmental intervention to minimize use of wood burning is urgently needed in south-central Chile and other high-income countries to provide health, environmental and economic co-benefits.

## Data Availability

All air pollution measurements from the beta attenuation monitors used in this study are publicly available (http://sinca.mma.gob.cl/) via the National Air Quality Information System (SINCA). Air pollution data obtained from the SPS30 sensors is publicly available from a mobile and web application (AIRE Temuco: http://aire.ceisufro.cl/#/dashboard). 

http://aire.ceisufro.cl/#/dashboard

## Acknowledgements

The authors would like to thank the AIRE Temuco team for their technical assistance with air monitor specifications and calibration methodology. The authors received no external sources of funding to conduct this work. Matthew Shupler is funded by a grant from the National Institute for Health Research (NIHR) (ref: 17/63/155) using UK aid from the UK government to support global health research. The views expressed in this publication are those of the authors and not necessarily those of the NIHR or the UK Department of Health and Social Care.

## References

Ahmed, S., Simiyu, E., Githiri, G., Sverdlik, A., Mbaka, S., 2015. Cooking up a storm Community-led mapping and. AIRE CEIUFRO, 2019.

Apte, J., Marshall, J., Cohen, A., Brauer, M., 2015. Addressing Global Mortality from Ambient PM2. 5 49, 8057–66. https://doi.org/10.1021/acs.est.5b01236

Barton, J.R., Ramírez, M.I., 2019. The Role of Planning Policies in Promoting Urban Sprawl in Intermediate Cities: Evidence from Chile. Sustainability (Switzerland) 11, 1–17. https://doi.org/10.3390/su11247165

Becerra, M, Jerez, A., Valenzuela, M., Garcés, H.O., Demarco, R., 2018. Life quality disparity: Analysis of indoor comfort gaps for Chilean households. Energy Policy 121, 190–201. https://doi.org/10.1016/j.enpol.2018.06.010

Becerra, Miguel, Jerez, A., Valenzuela, M., Garcés, H.O., Demarco, R., 2018. Life quality disparity: Analysis of indoor comfort gaps for Chilean households. Energy Policy 121, 190–201. https://doi.org/10.1016/j.enpol.2018.06.010

Bond, T.C., Doherty, S.J., Fahey, D.W., Forster, P.M., Berntsen, T., Deangelo, B.J., Flanner, M.G., Ghan, S., Kärcher, B., Koch, D., Kinne, S., Kondo, Y., Quinn, P.K., Sarofim, M.C., Schultz, M.G., Schulz, M., Venkataraman, C., Zhang, H., Zhang, S., Bellouin, N., Guttikunda, S.K., Hopke, P.K., Jacobson, M.Z., Kaiser, J.W., Klimont, Z., Lohmann, U., Schwarz, J.P., Shindell, D., Storelvmo, T., Warren, S.G., Zender, C.S., 2013. Bounding the role of black carbon in the climate system: A scientific assessment. Journal of Geophysical Research Atmospheres 118, 5380–5552. https://doi.org/10.1002/jgrd.50171

Boso, A., Hofflinger, A., Garrido, J., & Álvarez, B., 2020. Breathing clean air or cheaply heating your home: an environmental justice dilemma in Chilean Patagonia. Geographical Review 2507, 1–9.

Boso, Á., Hofflinger, A., Oltra, C., Alvarez, B., Garrido, J., 2018. Public support for wood smoke mitigation policies in south-central Chile. Air Qual Atmos Health 11, 1109–19.

Bravo-Linares, C, Ovando-Fuentealba, L., Orellana-Donoso, S., Gatica, S., Klerman, F M., Mudge, S., 2016. Source identification, apportionment and toxicity of indoor and outdoor PM 2. 5 airborne particulates in a region characterised by wood burning. Environmental Science: Processes & Impacts 18, 575–89.

Bravo-Linares, Claudio, Ovando-Fuentealba, L., Orellana-Donoso, S., Gatica, S., Klerman, F.M. Mudge, S., Gallardo, W., Paul Pinaud, J., Loyola-Sepulveda, R., 2016. Source identification, apportionment and toxicity of indoor and outdoor PM 2. 5 airborne particulates in a region characterised by wood burning. Environmental Science: Processes & Impacts 18, 575–589. https://doi.org/10.1039/C6EM00148C

British Standard Institution, 2008. BS EN ISO 15927-6:2007. Hygrothermal performance of buildings - Calculation and presentation of climatic data - Part 6: Accumulated temperature differences (degree-days) 22.

Bruce-Keller, A.J., Salbaum, J.M., Luo, M., Blanchard, E., Taylor, C.M., Welsh, D.A., Berthoud, H. -R., 2018. Low-cost sensors for the measurement of atmospheric composition: overview of topic and future applications, Wmo.

CDT, 2010. Estudio de usos finales y curva de oferta de la conservacion de la energía en el sector residencial. Santiago, Chile.

Chafe, Z.A., Brauer, M., Klimont, Z., Van Dingenen, R., Mehta, S., Rao, S., Riahi, K., Dentener, F., Smith, K.R., 2015. Household cooking with solid fuels contributes to ambient PM2. 5air pollution and the burden of disease. Environmental Health Perspectives 122, 1314–1320. https://doi.org/10.1289/ehp.1206340

Chauhan, A., Singh, R.P., 2020. Decline in PM 2. 5 concentrations over major cities around the world associated with COVID-19. Environmental Research 187, 109634. https://doi.org/10.1016/j.envres.2020.109634

Chen, Y., Senthilkumar, N., Shen, H., Shen, G., 2021. Environmental Inequality Deepened during the COVID-19 in the Developing World. Environmental Science and Technology 55, 7–8. https://doi.org/10.1021/acs.est.0c06193

Coccia, M., 2020. Factors determining the diffusion of COVID-19 and suggested strategy to prevent future accelerated viral infectivity similar to COVID. Science of The Total Environment 729, 138474. https://doi.org/10.1016/j.scitotenv.2020.138474

Collivignarelli, M. cristina, Abbà, A., Bertanza, G., Pedrazzani, R., Ricciardi, P., Carnevale, M., 2020. Lockdown for CoViD-2019 in Milan?: What are the effects on air quality?? Science of the Total Environment 732, 139280. https://doi.org/10.1016/j.scitotenv.2020.139280

Cuadrado, C., Monsalves, M.J., Gajardo, J., Bertoglia, M.P., Nájera, M., Alfaro, T., Canals, M., Kaufman, J.S., Peña, S., 2020. Impact of small-area lockdowns for the control of the COVID-19 pandemic. medRxiv. https://doi.org/10.1101/2020.05.05.20092106

Díaz-Robles, L., Díaz-Robles, L., Cortés, S., Carlos Ortega, J., Vergara-Fernández, A., 2015. Short term health effects of particulate matter: A comparison between wood smoke and multi-source polluted urban areas in chile. Aerosol and Air Quality Research 15, 306–318. https://doi.org/10.4209/aaqr.2013.10.0316

Díaz-Robles, L., Ortega, J.C., Fu, J.S., Reed, G.D., Chow, J.C., Watson, J.G., Moncada-Herrera, J.A., 2008. A hybrid ARIMA and artificial neural networks model to forecast particulate matter in urban areas: The case of Temuco, Chile. Atmospheric Environment 42, 8331–8340.

Díaz-Robles, L.A., Fu, J.S., Vergara-Fernández, A., Etcharren, P., Schiappacasse, L.N., Reed, G.D., Silva, M.P., 2014. Health risks caused by short term exposure to ultrafine particles generated by residential wood combustion: A case study of Temuco, Chile. Environment International 66, 174–181.

Dickson, R.R., 1961. Meteorological Factors Affecting Particulate Air Pollution of a City. Bulletin of the American Meteorological Society 42, 556–560. https://doi.org/10.1175/1520-0477-42.8.556

Dirección Meteorológica de Chile – Servicios Climáticos, Dirección General De Aeronáutica Civil (DGAC). (https://climatologia.meteochile.gob.cl/)., n. d.

Evangeliou, N., Platt, S., Eckhardt, S., Lund Myhre, C., Laj, P., Alados-Arboledas, L., Backman, J., Brem, B., Fiebig, M., Flentje, H., Marinoni, A., Pandolfi, M., Yus-Dìez, J., Prats, N., Putaud, J., Sellegri, K., Sorribas, M., Eleftheriadis, K., Vratolis, S., Wiedensohler, A., Stohl, A., 2020. Changes in black carbon emissions over Europe due to COVID-19 lockdowns. Atmospheric Chemistry and Physics 1–33. https://doi.org/10.5194/acp-2020-1005

Fattorini, D., Regoli, F., 2020. Role of the atmospheric pollution in the Covid-19 outbreak risk in Italy. medRxiv 264, 2020.04.23.20076455. https://doi.org/10.1101/2020.04.23.20076455

Garín Contreras, A., Salvo Garrido, S., Bravo Araneda, G., 2009. Segregación residencial y políticas de vivienda en Temuco: 1992-2002. Revista de geografía Norte Grande. https://doi.org/10.4067/s0718-34022009000300006

Gayen, A., Haque, S.M., Mishra, S.V., 2021. COVID-19 induced lockdown and decreasing particulate matter (PM10): An empirical investigation of an Asian megacity. Urban Climate 36, 33552884. https://doi.org/10.1016/j.uclim.2021.100786

Gobeli, D., Schloesser, H., Pottberg, T., 2008. Met one instruments BAM-1020 beta attenuation mass monitor US-EPA PM2. 5 federal equivalent method field test results The Air & Waste Management Association (A&WMA). Conference, Kansas City, Mo 2008.

Gozzi, N., Tizzoni, M., Chinazzi, M., Ferres, L., Vespignani, A., Perra, N., 2020. Estimating the effect of social inequalities in the mitigation of COVID-19 across communities in Santiago de Chile. medRxiv 1–19. https://doi.org/10.1101/2020.10.08.20204750

Hasnain, M., Pasha, M.F., Ghani, I., 2020. Combined Measures to Control the COVID-19 Pandemic in Wuhan, Hubei, China: A Narrative Review. Journal of Biosafety and Biosecurity 2, 51–57. https://doi.org/10.1016/j.jobb.2020.10.001

Howden-Chapman, P., Chapman, R., 2012. Health co-benefits from housing-related policies. Current Opinion in Environmental Sustainability 4, 414–419. https://doi.org/10.1016/j.cosust.2012.08.010

Huang, Y., Partha, D.B., Harper, K., Heyes, C., 2021. Impacts of global solid biofuel stove emissions on ambient air quality and human health. GeoHealth 1–16. https://doi.org/10.1029/2020gh000362

IBM Corp, 2017. IBM SPSS Statistics for Windows. IBM Corp, Armonk, NY.

Jay, J., Bor, J., Nsoesie, E.O., Lipson, S.K., Jones, D.K., Galea, S., Raifman, J., 2020. Neighbourhood income and physical distancing during the COVID-19 pandemic in the United States. Nature Human Behaviour 4, 1294–1302. https://doi.org/10.1038/s41562-020-00998-2

Jorquera, H., Barraza, F., Heyer, J., Valdivia, G., Schiappacasse, L., Ld, M., 2018. Indoor PM2. 5 in an urban zone with heavy wood smoke pollution: The case of Temuco, Chile. Environmental Pollution 236, 477–487. https://doi.org/10.1016/j.envpol.2018.01.085.

Kumar, P., Morawska, L., Martani, C., Biskos, G., Neophytou, M., Di Sabatino, S., Bell, M., Norford, L., Britter, R., 2015. The rise of low-cost sensing for managing air pollution in cities. Environment International 75, 199–205. https://doi.org/10.1016/j.envint.2014.11.019

Lai, A.M., Ellison Carter, Shan, M., Ni, K., Clark, S., Ezzati, M., Wiedinmyer, C., Yang, X., Baumgartner, J., Schauer, J.J., 2019. Chemical composition and source apportionment of ambient, household, and personal exposures to PM2. 5 in communities using biomass stoves in rural China. Science of the Total Environment 646, 309–319.

Liddell, C., Morris, C., Thomson, H., Guiney, C., 2016. Excess winter deaths in 30 European countries 1980-2013: A critical review of methods. Journal of Public Health (United Kingdom) 38, 806–814. https://doi.org/10.1093/pubmed/fdv184

Maag, B., Zhou, Z., Thiele, L., 2018. A Survey on Sensor Calibration in Air Pollution Monitoring Deployments.

IEEE Internet of Things Journal 5, 4857–4870. https://doi.org/10.1109/JIOT.2018.2853660

Mahato, S., Pal, S., Ghosh, K.G., 2020. Effect of lockdown amid COVID-19 pandemic on air quality of the megacity Delhi, India. Science of The Total Environment 730, 139086. https://doi.org/10.1016/j.scitotenv.2020.139086

Malings, C., Tanzer, R., Hauryliuk, A., Saha, P.K., Robinson, A.L., Presto, A.A., Subramanian, R., 2020. Fine particle mass monitoring with low-cost sensors: Corrections and long-term performance evaluation. Aerosol Science and Technology 54, 160–174. https://doi.org/10.1080/02786826.2019.1623863

Mardones, C., Cornejo, N., 2020. Ex-post evaluation of environmental decontamination plans on air quality in Chilean cities. Journal of Environmental Management 256, 109929. https://doi.org/10.1016/j.jenvman.2019.109929.

Martinez-Soto, A., Jentsch, M.F., 2019. A transferable energy model for determining the future energy demand and its uncertainty in a country’s residential sector. Building Research & Information 1–26. https://doi.org/10.1080/09613218.2019.1692188

Mena, G., Martinez, P.P., Mahmud, A.S., Marquet, P.A., Buckee, C.O., Santillana, M., 2021. Socioeconomic status determines COVID-19 incidence and related mortality in Santiago, Chile. medRxiv 2021.01. 12.21249682.

Ministerio de Desarrollo Social [MIDESO], 2018. INGRESOS DE LOS HOGARES. Síntesis de resultados Casen 2017 72.

MMA, 2018. DS N°48 Plan de descontaminación atmosférica por MP 2, 5 para las comuna de Temuco y Padre las casas actualización del plan de descontaminación por MP 10, para las mismas comunas DS N°8 2018.

Molina, C., Toro A, R., Morales S, R.G.E., Manzano, C., Leiva-Guzmán, M.A., 2017. Particulate matter in urban areas of south-central Chile exceeds air quality standards. Air Qual Atmos Health 10, 653–667. https://doi.org/10.1007/s11869-017-0459-y

Murray, C., Abbafati, C., Machado, D.B., Cislaghi, B., Salman, O.M., Karanikolos, M., McKee, M., Abbas, K.M., Brady, O.J., Larson, H.J., Trias-Llimós, S., Cummins, S., Langan, S.M., Sartorius, B., Hafiz, A., Jenabi, E., Mohammad Gholi Mezerji, N., Borzouei, S., Azarian, G., Khazaei, S., Abbasi, M., Asghari, B., Masoumi, S., Komaki, H., Taherkhani, A., Adabi, M., Abbasifard, M., Bazmandegan, G., Kamiab, Z., Vakilian, A., Anjomshoa, M., Mokari, A., Sabour, S., Shahbaz, M., Saeedi, R., Ahmadieh, H., Yousefinezhadi, T., Haj-Mirzaian, A., Nikbakhsh, R., Safi, S., Asgari, S., Irvani, S.N., Jahanmehr, N., Ramezanzadeh, K., Abbasi-Kangevari, M., Khayamzadeh, M., Abbastabar, H., Shirkoohi, R., Fazlzadeh, M., Janjani, H., Hosseini, M., Mansournia, M., Tohidinik, H., Bakhtiari, A., Fazaeli, A., Mousavi, S., Hasanzadeh, A., Nabavizadeh, B., Malekzadeh, R., Hashemian, M., Pourshams, A., Salimzadeh, H., Sepanlou, S.G., Afarideh, M., Esteghamati, A., Esteghamati, S., Ghajar, A., Heidari, B., Rezaei, N., Mohamadi, E., Rahimi-Movaghar, A., Rahim, F., Eskandarieh, S., Sahraian, M., Mohebi, F., Aminorroaya, A., Ebrahimi, H., Farzadfar, F., Mohajer, B., Pishgar, F., Saeedi Moghaddam, S., Shabani, M., Zarafshan, H., Abolhassani, H., Hafezi-Nejad, N., Heidari-Soureshjani, R., Abdollahi, M., Farahmand, M., Salamati, P., Mehrabi Nasab, E., Tajdini, M., Aghamir, S., Mirzaei, R., Dibaji Forooshani, Z., Khater, M.M., Abd-Allah, F., Abdelalim, A., Abualhasan, A., El-Jaafary, S.I., Hassan, A., Elsharkawy, A., Khater, A.M., Elhabashy, H.R., Salem, M.R.R., Salem, H., Sadeghi, M., Jafarinia, M., Amini-Rarani, M., Mohammadifard, N., Sarrafzadegan, N., Abdollahpour, I., Sarveazad, A., Tehrani-Banihashemi, A., Yoosefi Lebni, J., Manafi, N., Pazoki Toroudi, H., Dorostkar, F., Alipour, V., Sheikhtaheri, A., Arabloo, J., Azari, S., Ghashghaee, A., Rezapour, A., Naserbakht, M., Kabir, A., Mehri, F., Yousefifard, M., Asadi-Aliabadi, M., Babaee, E., Eshrati, B., Goharinezhad, S., Moradi, M., Abedi, P., Rashedi, V., Kumar, V., Elgendy, I.Y., Basu, S., Park, J., Pereira, A., Norheim, O.F., Eagan, A.W., Cahill, L.E., Sheikh, A., Abushouk, A.I., Kraemer, M.U.G., Thakur, B., Bärnighausen, T.W., Shrime, M.G., Abedi, A., Doshi, C.P., Abegaz, K.H., Geberemariyam, B.S., Aynalem, Y.A., Shiferaw, W.S., Abosetugn, A.E., Aboyans, V., Abrams, E.M., Gitimoghaddam, M., Kissoon, N., Stubbs, J.L., Brauer, M., Iyamu, I.O., Kopec, J.A., Pourmalek, F., Ribeiro, A.P., Malta, D.C., Gomez, R.S., Abreu, L.G., Abrigo, M.R.M., Almulhim, A.M., Dahlawi, S.M.A., Pottoo, F.H., Menezes, R.G., Alanzi, T.M., Alumran, A.K., Abu Haimed, A.K., Madadin, M., Alanezi, F.M., Abu-Gharbieh, E., Saddik, B., Abu-Raddad, L.J., Samy, A.M., El Nahas, N., Shalash, A.S., Nabhan, A.F., Kamath, A.M., Kassebaum, N.J., Aravkin, A.Y., Kochhar, S., Sorensen, R.J.D., Afshin, A., Burkart, K., Cromwell, E.A., Dandona, L., Dharmaratne, S.D., Gakidou, E., Hay, S.I., Kyu, H.H., Lopez, A.D., Lozano, R., Misganaw, A.T., Mokdad, A.H., Naghavi, M., Pigott, D.M., Reiner, R.C., Roth, G.A., Stanaway, J.D., Vollset, S., Vos, T., Wang, H., Lim, S.S., Murray, C.J.L., Kalani, R., Ikuta, K.S., Cho, D.Y., Kneib, C.J., Crowe, C.S., Massenburg, B.B., Morrison, S.D., Acebedo, A., Adelson, J.D., Agesa, K.M., Alam, T., Albertson, S.B., Anderson, J.A., Antony, C.M., Ashbaugh, C., Assmus, M., Azhar, G., Balassyano, S., Bannick, M.S., Barthelemy, C.M., Bender, R.G., Bennitt, F.B., Bertolacci, G.J., Biehl, M.H., Bisignano, C., Boon-Dooley, A.S., Briant, P.S., Bryazka, D., Bumgarner, B.R., Callender, C.S., Cao, J., Castle, C.D., Castro, E., Causey, K., Cercy, K.M., Chalek, J., Charlson, F.J., Cohen, A.J., Comfort, H., Compton, K., Croneberger, A.J., Cruz, J.A., Cunningham, M., Dandona, R., Dangel, W.J., Dean, F.E., DeCleene, N.K., Deen, A., Degenhardt, L., Dingels, Z.V., Dippenaar, I.N., Dirac, M.A., Dolgert, A.J., Emmons-Bell, S., Estep, K., Farag, T., Feigin, V.L., Feldman, R., Ferrara, G., Ferrari, A.J., Fitzgerald, R., Force, L.M., Fox, J.T., Frank, T.D., Fu, W., Fukutaki, K., Fuller, J.E., Fullman, N., Galles, N.C., Gardner, W.M., Gershberg Hayoon, A., Goren, E., Gorman, T.M., Gottlich, H.C., Guo, G., Haddock, B., Hagins, H., Haile, L.M., Hamilton, E.B., Han, C., Han, H., Harvey, J.D., Henny, K., Henrikson, H.J., Henry, N.J., Herbert, M.E., Hsiao, T., Huynh, C.K., Iannucci, V.C., Ippolito, H., Irvine, C.M.S., Jafari, H., Jahagirdar, D., James, S.L., Johnson, C.O., Johnson, S.C., Keller, C., Kemmer, L., Kendrick, P.J., Knight, M., Kocarnik, J.M., Krohn, K.J., Larson, S.L., Lau, K.M., Ledesma, J.R., Leever, A.T., LeGrand, K.E., Lescinsky, H., Lin, C., Liu, H., Liu, Z., Lo, J., Lu, A., Ma, J., Maddison, E.R., Manguerra, H., Marks, A., Martopullo, I., Mastrogiacomo, C.I., May, E.A., Mooney, M.D., Mosser, J.F., Mullany, E.C., Mumford, J., Munro, S.B., Nandakumar, V., Nguyen, J., Nguyen, M., Nichols, E., Nixon, M.R., Odell, C.M., Ong, K.L., Orji, A.U., Ostroff, S.M., Pasovic, M., Paulson, K.R., Pease, S.A., Pennini, A., Pierce, M., Pilz, T.M., Pletcher, M., Rao, P.C., Razo, C., Redford, S.B., Reinig, N., Reitsma, M.B., Rhinehart, P., Robalik, T., Roberts, S., Roberts, N.L.S., Rolfe, S., Sbarra, A.N., Schaeffer, L.E., Shackelford, K.A., Shadid, J., Sharara, F., Shaw, D.H., Sheena, B.S., Simpson, K.E., Smith, A., Spencer, C.N., Spurlock, E.E., Stark, B.A., Steiner, C., Steuben, K.M., Sylte, D.O., Tang, M., Taylor, H.J., Terrason, S., Thomson, A.M., Torre, A.E., Travillian, R., Troeger, C.E., Vongpradith, A., Walters, M.K., Wang, J., Watson, A., Watson, S., Whisnant, J.L., Whiteford, H.A., Wiens, K.E., Wilner, L.B., Wilson, S., Wool, E.E., Wozniak, S.S., Wu, J., Wulf Hanson, S., Wunrow, H., Xu, R., Yadgir, S., Yearwood, J.A., York, H.W., Yuan, C., Zhao, J.T., Zheng, P., Zimsen, S.R.M., Zlavog, B.S., Chang, A.Y., Oren, E., Buchbinder, R., Chin, K.L., Guo, Y., Polkinghorne, K.R., Thrift, A.G., Lee, S.W.H., Ackerman, I.N., Cicuttini, F.M., Li, S., Zaman, S., Suleria, H., Zhang, J., Cowie, B.C., Wijeratne, T., Patton, G.C., Sawyer, S.M., Adair, T., Meretoja, A., Adetokunboh, O.O., Adamu, A.A., Iwu, C.J., Parry, C.D.H., Seedat, S., Ndwandwe, D.E., Mahasha, P.W., Stein, D.J., Nnaji, C.A., Sambala, E.Z., Wiysonge, C.S., Adebayo, O.M., Ilesanmi, O.S., Owolabi, M.O., Adeoye, A.M., Adedeji, I.A., Adekanmbi, V., Ibitoye, S.E., John-Akinola, Y.O., Oluwasanu, M.M., Oghenetega, O.B., Akinyemi, R.O., Zandian, H., Adham, D., Zahirian Moghadam, T., Advani, S.M., Teagle, W.L., Braithwaite, D., Agasthi, P., Saadatagah, S., Afshari, M., Agardh, E.E., Allebeck, P., Danielsson, A., Deuba, K., Carrero, J.J., Mohammad, D.K., Fereshtehnejad, S., Ärnlöv, J., Nowak, C., Cederroth, C.R., Ahmadi, A., Pathak, A., Mills, E.J., Kurmi, O.P., Olagunju, A.T., Agarwal, G., Sathish, T., Aghaali, M., Mohammadbeigi, A., Agrawal, A., Ahmad, T., Ahmadi, K., Maleki, S., Naderi, M., Salahshoor, M.R., Pourmirza Kalhori, R., Almasi, A., Salimi, Y., Siabani, S., Ziapour, A., Barzegar, A., Khazaie, H., Kianipour, N., Amiri, F., Salehi Zahabi, S., Mirzaei, M., Shamsi, M., Najafi, F., Jalali, A., Ghadiri, K., Heydarpour, F., Fattahi, N., Karami Matin, B., Kazemi Karyani, A., Pirsaheb, M., Rajati, F., Sadeghi, E., Safari, Y., Sharafi, K., Soltani, S., Vasseghian, Y., Atafar, Z., Jalilian, F., Mirzaei-Alavijeh, M., Saeidi, S., Soofi, M., Zangeneh, A., Mansouri, B., Ahmadi, M., Khafaie, M.A., Safiri, S., Moghadaszadeh, M., Asghari Jafarabadi, M., Doshmangir, L., Jadidi-Niaragh, F., Ghafourifard, M., Spotin, A., Khodayari, M., Samadi Kafil, H., Kalankesh, L.R., Ahmadpour, E., Yousefi, B., Ansari, F., Hassankhani, H., Karimi, S., Haririan, H., Mereta, S., Ahmed, M.B., Feyissa, G.T., Ciobanu, L.G., Aji, B., Aynalem, G.L., Gebresillassie, B., Tefera, Y.G., Akalu, T.Y., Baraki, A.G., Tesema, G.A., Tessema, Z.T., Tamiru, A.T., Azene, Z.N., Netsere, H.B., Yano, Y., Akinyemiju, T., Wu, C., Zadey, S., Samad, Z., Ji, J.S., Doshi, P.P., John, O., Jha, V., Maulik, P.K., Pesudovs, K., Resnikoff, S., Mitchell, P.B., Sachdev, P.S., Akombi, B., Godinho, M.A., Ivers, R.Q., Peden, A.E., Biswas, R., Boufous, S., Akunna, C.J., Alahdab, F., Hammer, M.S., van Donkelaar, A., Al-Aly, Z., Dellavalle, R.P., Alam, S., Martin, R.V., Alam, N., De Leo, D., Tadakamadla, S.K., Alam, K., Alcalde-Rabanal, J.E., Avila-Burgos, L., Serván-Mori, E., Denova-Gutiérrez, E., Rodríguez-Ramírez, S., Morales, L., Poznańska, A., Wojtyniak, B., Rivera, J.A., Campos-Nonato, I.R., Campuzano Rincon, J., Sánchez-Pimienta, T.G., Mengesha, M.B., Welay, F.T., Alema, N.M., Demsie, D.G., Teame, H., Teklehaimanot, B.F., Alemu, B.W., Gultie, T., Bante, A.B., Yeshitila, Y.G., Geramo, Y.C.D., Glagn, M., Sorrie, M.B., W/hawariat, F.G., Amare, A., Kasa, A., Alemu, Y., Mihretie, K.M., Atnafu, D.D., Demeke, F.M., Melese, A., Bante, S.A., Dessie, G.A., Mengesha, E.W., Nigatu, D., Almadi, M.A.H., Alhabib, K.F., Mohammad, Y., Altirkawi, K.A., Temsah, M., Kugbey, N., Ayanore, M.A., Alhassan, R.K., Ali, M., Ali, S., Alicandro, G., Kalhor, R., Alijanzadeh, M., Alinia, C., Yusefzadeh, H., Didarloo, A., Alizade, H., Nikpoor, A., Aljunid, S.M., Alla, F., Leal, L.F., Almasi-Hashiani, A., Moradzadeh, R., Zamanian, M., Nazari, J., Amini, S., Ghamari, F., Almasri, N.A., Khan, M., Al-Mekhlafi, H.M., Bedi, N., Alonso, J., Ciber, E.S.P., Al-Raddadi, R.M., Zakzuk, J., Alvis-Guzman, N., Alvis-Zakzuk, N.J., Castañeda-Orjuela, C.A., Malagón-Rojas, J.N., Gezae, K., Gesesew, H.A., Muthupandian, S., Gebremeskel, G.G., Berhe, K., Amare, B., Ayza, M.A., Gebremeskel, L.G., Gebreslassie, A.A.A., Bitew, H., Zewdie, K.A., Tela, F.G.G., Gill, T.K., Noubiap, J., Lassi, Z.S., Bhandari, D., Amit, A.L., Antonio, C.T., Faraon, E.A., Lopez, J.F., Atre, S.R., Ballew, S.H., Matsushita, K., Khoja, A.T., Daneshpajouhnejad, P., Ghadimi, M., Shafaat, O., Fanzo, J., Amugsi, D.A., Amul, G.H., Koh, D.S.Q., Venketasubramanian, N., Anbesu, E.W., Mohammed, J.A., Wondmeneh, T.G., Andrei, C., Negoi, R.I., Davitoiu, D.V., Manda, A., Negoi, I., Preotescu, L., Hostiuc, M., Hostiuc, S., Ancuceanu, R., Hasan, M., Leung, J., Anderlini, D., Mamun, A.A., Maravilla, J.C., McGrath, J.J., Charlson, F.J., Lalloo, R., Uddin, R., Erskine, H.E., Knibbs, L.D., Mantilla Herrera, A. M., Santomauro, D.F., Mirica, A., Ausloos, M., Herteliu, C., Oţoiu, A., Pana, A., Andrei, T., Ştefan, S., Androudi, S., Angus, C., Ansari, I., Pourjafar, H., Shams-Beyranvand, M., Ansari-Moghaddam, A., Khammarnia, M., Antonazzo, I., Ferrara, P., Conti, S., Cortesi, P.A., Fornari, C., Lee, P.H., Antriyandarti, E., Yousefi, Z., Rafiei, A., Javidnia, J., Faridnia, R., Anvari, D., Goudarzi, H., Moosazadeh, M., Rezai, M., Daryani, A., Fareed, M., Anwer, R., Appiah, S., Paudel, D., Dichgans, M., Riahi, S., Rajabpour-Sanati, A., Arab-Zozani, M., Arba, A.A.K., Aremu, O., Ariani, F., Aripov, T., Armoon, B., Mahdavi, M.M., Arowosegbe, O.O., Tediosi, F., Aryal, K.K., Mosapour, A., Yaminfirooz, M., Arzani, A., Bijani, A., Jahani, M.A., Mouodi, S., Zamani, M., Asaad, M., Dianatinasab, M., Bahrami, M., Pakshir, K., Asadi-Pooya, A.A., Bayati, M., Shahabi, S., Athari, S., Atout, M.M.W., Atteraya, M.S., Brugha, T., Ausloos, F., Avokpaho, E.F.G., Room, R., Islam, M., Edvardsson, D., Rahman, M., Ayala Quintanilla, B., Gething, P.W., Briggs, A.M., Ayano, G., Hendrie, D., Miller, T.R., Azzopardi, P.S., Hoogar, P., B, D. B., Kulkarni, V., Kumar, N., Mithra, P., Shetty, R.S., Thapar, R., Padubidri, J., Bakkannavar, S.M., Nayak, V.C., Rastogi, P., Shetty, B.K., Bhageerathy, R., Gudi, N., Boloor, A., Holla, R., Rathi, P., Unnikrishnan, B., Janodia, M.D., Lang, J.J., Badawi, A., Orpana, H.M., Bhutta, Z.A., Shield, K.D., Chattu, V., Badiye, A.D., Kapoor, N., Bagherzadeh, M., Rabiee, N., Bagli, E., Baig, A.A., Bairwa, M., Lodha, R., Sagar, R., Rath, G.K., Bhardwaj, P., Charan, J., Kanchan, T., Joshi, A., Pakhare, A.P., Bakhshaei, M., Naghshtabrizi, B., Balachandran, A., Hoek, H.W., Postma, M.J., Geremew, A., Gebrehiwot, A.M., Balakrishnan, S., Desalew, A., Bojia, H.A., Mohammed, A.S., Regassa, L.D., Parmar, P.G.K., Balalla, S., Roberts, S., Baldasseroni, A., Stokes, M.A., Ball, K., Islam, S., Maddison, R., Balzi, D., Levi, M., Banach, M., Banerjee, S.K., Banik, P.C., Barua, L., Faruque, M., Barboza, M.A., Barker-Collo, S.L., Jonas, J.B., Panda-Jonas, S., De Neve, J., Kohler, S., Moazen, B., Mohammed, S., Barrero, L.H., Basaleem, H., Bassat, Q., Haro, J.M., Koyanagi, A., Car, J., Greaves, F., Majeed, A., Davis, A.C., Steiner, T.J., Kusuma, D., Palladino, R., Rawaf, S., Saxena, S., Rawaf, D.L., Baune, B.T., Karch, A., Baye, B.A., Darega Gela, J., Kolola, T., Becker, J.S., DeLang, M., West, J., Gad, M.M., Serre, M.L., Gallus, S., Lugo, A., Beghi, E., Pupillo, E., Bosetti, C., Giussani, G., Bikbov, B., Perico, N., Remuzzi, G., Imani-Nasab, M., Nouraei Motlagh, S., Sharafi, Z., Behzadifar, M., Béjot, Y., Bekuma, T.T., Yilma, M.T., Bell, M.L., Bello, A.K., Rafiee, A., Keddie, S.H., Lucas, T.C.D., Dolecek, C., Dunachie, S.J., Kraemer, M.U.G., Rumisha, S.F., Weiss, D.J., Lewington, S., Collins, E.L., Nandi, A.K., Zhao, Y., Bennett, D.A., Karim, M.A., Lacey, B., Khundkar, R., Yaya, S., Goulart, A.C., Santos, I.S., Bensenor, I.M., Lotufo, P.A., Tovani-Palone, M.R., Castaldelli-Maia, J., Wang, Y., Furtado, J.M., Benziger, C.P., Berman, A.E., Mazidi, M., Bernabe, E., Dargan, P.I., Molokhia, M., Shibuya, K., Douiri, A., Wolfe, C.D.A., Hay, R.J., Flohr, C., Suchdev, P.S., Ram, P., Bernstein, R.S., Liu, Y., Bhagavathula, A.S., Khan, G., Grivna, M., Bhala, N., Chandan, J.S., Gaidhane, A.M., Quazi Syed, Z., Saxena, D., Khatib, M., Bhat, A.G., Bhattacharyya, K., Bhattarai, S., Das, J.K., Bibi, S., Bilano, V., Bin Sayeed, M., Cherbuin, N., D’Amico, E., Grosso, G., Borzì, A.M., Biondi, A., Vacante, M., Valli, A., Birihane, B.M., Bisanzio, D., Hassan, S., Bjørge, T., Øverland, S., Bockarie, M.J., Mensah, G.A., Sliwa, K., Gholamian, A., Bohlouli, S., Esmaeilnejad, S., Bohluli, M., Bolla, S.R., Borges, G., Bose, D., Bourne, R., Brayne, C., Breitborde, N.J.K., Fisher, J.L., Breitner, S., Brenner, H., Breusov, A.V., Rakovac, I., Briko, N.I., Lopukhov, P.D., Glushkova, E.V., Korshunov, V.A., Polibin, R.V., Jakovljevic, M., Briko, A.N., Britton, G.B., Castro, F., Moreno Velásquez, I., Burnett, R.T., Burugina Nagaraja, S., Busse, R., Butt, Z.A., Caetano dos Santos, F., Cai, T., Hall, B.J., Cahuana-Hurtado, L., Cámera, L.A., Valdez, P.R., Tudor Car, L., Cárdenas, R., Gorini, G., Carreras, G., Fernandes, E., Freitas, M., Pereira, D.M., Santos, J.V., Ribeiro, D., Pinheiro, M., Massano, J., das Neves, J., Dias da Silva, D., Carvalho, F., Costa, V.M., Silva, J.P., Morgado-da-Costa, J., Castelpietra, G., Catalá-López, F., Pakhale, S., Cerin, E., Yip, P., Ho, H., Chang, J., Chang, A.R., Chang, K., Cooper, O.R., Chaturvedi, S., Chimed-Ochir, O., Chirinos-Caceres, J.L., Choi, J.J., Christensen, H., Truelsen, T.C., Chu, D., Kivimäki, M., Chung, S., Kumar, M., Ward, J.L., Chung, M.T., Cirillo, M., Avic, S., Classen, T.K.D., Lauriola, P., Corso, B., Hugo, F.N., Kieling, C., Cousin, E., Duncan, B.B., Goulart, B.N.G., Schmidt, M.I., Stein, C., Cowden, R.G., MacLachlan, J.H., Leigh, J., Cross, M., Ferreira, M.L., Smith, E.U.R., Driscoll, T.R., Huda, T.M., Hoy, D.G., Horita, N., Cross, D.H., Dai, H., Hu, G., Damasceno, A.A.M., Damiani, G., La Vecchia, C., Sanabria, J., Pandey, A., Mathur, M.R., Zodpey, S., Dandona, L., Dandona, R., Kumar, G., Lal, D.K., Hamagharib Abdullah, K., Hosseinzadeh, M., Darwesh, A.M., Faraj, A., Omar Bali, A., Das Gupta, R., Dash, A.P., Davey, G., Deribe, K., Gebremedhin, K.B., Wondmieneh, A.B., Dereje, N.D., Dávila-Cervantes, C.A., Davletov, K., Mereke, A., Serdar, B., Gebremeskel, G.G., Haile, T.G., Tadesse, D.B., Weldesamuel, G.T., Demoz, G.T., Gebremeskel, L.G., Woldu, G., Handiso, D., Dervenis, N., Topouzis, F., Desai, R., Dharmaratne, S.D., Dhungana, G.P., Emamian, M., Diaz, D., Djalalinia, S., Nguyen, T.H., Vu, G.T., Do, H.T., Dokova, K., Doku, D.T., Neupane, S., Takala, J.S., Doxey, M.C., Doyle, K.E., Renzaho, A.M.N., Ogbo, F.A., Robinson, S.R., Vukovic, A., Ilic, I.M., Santric-Milicevic, M.M., Vujcic, I.S., Dubljanin, E., Vukovic, R., Rasella, D., Duraes, A.R., Ebrahimi Kalan, M., Effiong, A., Pond, C.D., Ehrlich, J.R., Liu, X., Wei, M.Y.W., El Sayed, I., El Tantawi, M., El Sayed Zaki, M., Elbarazi, I., Tsai, A.C., Elyazar, I.R., Yazdi-Feyzabadi, V., Eskandari, K., Sharifi, H., Halvaei, I., Ghaffarifar, F., Zaki, L., Hasanpoor, E., Esmaeilzadeh, F., Etemadi, A., Lan, Q., Yeheyis, T.Y., Etisso, A.E., Ezekannagha, O., e Farinha, C.S., Farioli, A., Violante, F.S., Faris, P.S., Mohammad, D.K., Faro, A., Lam, J.O., Filip, I., Fischer, F., Flor, L.S., Foigt, N.A., Folayan, M.O., Fomenkov, A.A., Foroutan, M., Francis, J.M., Franklin, R.C., Fukumoto, T., Gamkrelidze, A., Kereselidze, M., Garcia-Basteiro, A.L., Lazarus, J.V., Straif, K., Ghiasvand, H., Ghith, N., Norrving, B., Mehta, K.M., Mohamad, O., Salama, J.S., Ghosh, R., Giampaoli, S., Gilani, S., Hameed, S., Hanif, A., Rana, S.M., Uthman, O.A., Gill, P.S., Gillum, R.F., Ginawi, I.A., Ginindza, T.G., Tlou, B., Tanser, F.C., Gnedovskaya, E.V., Kravchenko, M.A., Piradov, M.A., Golechha, M., Goli, S., Milne, G.J., Schlaich, M.P., Hankey, G.J., Gona, P.N., Gopalani, S.V., Goudarzi, H., Rawassizadeh, R., Grada, A., Nsoesie, E.O., Javaheri, T., Rawaf, S., Steel, N., Gubari, M.I.M., Gugnani, H.C., Guimaraes, A.L.S., Guimarães, R.A., Guled, R.A., Senbeta, A.M., Hashi, A., Omer, M.O., Yousuf, A.Y., Gupta, S.S., Gupta, T., Gupta, R., Gupta, R., Haagsma, J.A., Polinder, S., Hachinski, V., Stranges, S., Hamadeh, R.R., Hamidi, S., Tabarés-Seisdedos, R., Tyrovolas, S., Hasaballah, A.I., Hassanipour, S., Joukar, F., Mansour-Ghanaei, F., Jaafari, J., Havmoeller, R.J., Hayat, K., Heibati, B., Heidari, G., Henok, A., Hird, T.R., Moraga, P., Sigfusdottir, I.D., Phillips, M.R., Hole, M.K., Hollingsworth, B., Hopf, K.P., Hosgood, H., Hossain, N., Mai, H.T., Nguyen, C.T., Nguyen, D.N., Nguyen, H.L.T., Pham, H.Q., Nguyen, D.N., Househ, M., Hsairi, M., Lin, R., Hsieh, V., Hwang, B., Hu, K., Humayun, A., Hussain, R., Ibeneme, C.U., Ikeda, N., Ilic, M.D., Inbaraj, L.R., Iqbal, U., Islami, F., Yamagishi, K., Iso, H., Iwu, C.C.D., Jacobsen, K.H., Jayatilleke, A.U., Jeemon, P., Jha, R.P., Jia, P., Osei, F.B., Schutte, A.E., Johansson, L., Szócska, M., Joo, T., Jozwiak, J.J., Jürisson, M., Kabir, Z., Roshandel, G., Oladnabi, M., Kalani, H., Kassa, G.M., Katikireddi, S.V., Yamada, T., Nomura, S., Kawakami, N., Kayode, G.A., Kromhout, H., Traini, E., Kumar, M., Keiyoro, P.N., Muriithi, M.K., Wamai, R.G., Khader, Y.S., Khalid, N., Khan, E.A., Khang, Y., Khatab, K., Khubchandani, J., Kim, Y., Yoon, S., Shin, M., Kim, C., Kim, Y., Kim, D., Kimokoti, R.W., Kinfu, Y., Kisa, S., Kisa, A., Kissimova-Skarbek, K., Topor-Madry, R., Lallukka, T., Meretoja, T.J., Shivakumar, K.M., Knudsen, A.S., Sulo, G., Kosen, S., Kotlo, A., Koul, P.A., Krishan, K., Ks, S., Kuate Defo, B., Kucuk Bicer, B., Yuce, D., Rawat, R., Meitei, W.B., Kumar, P., Singh, P., Kumaresh, G., Lami, F.H., Landires, I., Nunez-Samudio, V., Landires, Lansingh V. C., Lansky, S., Larsson, A.O., Lasrado, S., Lavados, P.M., Leasher, J.L., Lee, S.W.H., Leonardi, M., Raggi, A., Sattin PsyD, S Schiavolin, D., Li, B., Lim, L., Takahashi, K., Linehan, C., Linn, S., Shuval, K., Listl, S., Zhou, M., Liu, S., Logroscino, G., Looker, K.J., Lorkowski, S., Lunevicius, R., Lyons, R.A., Madotto, F., Magdy Abd El Razek, H., Magdy Abd El Razek, M., Mahmoudi, M., Maled, V., Moradi, G., Maleki, A., Manafi, A., Mapoma, C., Martini, S., Martins-Melo, F.R., Masaka, A., McAlinden, C., Momen, N.C., Plana-Ripoll, O., Medina-Solís, C.E., Meharie, B., Mehndiratta, M., Mehrotra, R., Suliankatchi Abdulkader, R., Mekonnen, T., Memiah, P.T.N., Wallin, M.T., Memish, Z.A., Mendoza, W., Mestrovic, T., Miazgowski, B., Miazgowski, T., Miazgowski, B., Michalek, I., Mini, G., Miri, M., Mirrakhimov, E.M., Mirzaei, H., Mohammadian-Hafshejani, A., Mohammadpourhodki, R., Sahebkar, A., Mohammed, H., Sufiyan, M.B., Mohammed, S., Mohseni Bandpei, M. A., Monasta, L., Ronfani, L., Mondello, S., Moradi-Joo, M., Morawska, L., Mousavi Khaneghah, A., Westerman, R., Werdecker, A., Mueller, U.O., Mukhopadhyay, S., Musa, K., Mustafa, G., Nagarajan, A.J., Nagel, G., Rothenbacher, D., Naik, G., Schwebel, D.C., Singh, J.A., Naimzada, M., Otstavnov, N., Otstavnov, S.S., Soshnikov, S., Titova, M.V., Nair, S., Naldi, L., Nangia, V., Nansseu, J., Sobngwi, E., Nguefack-Tsague, G., Ndejjo, R., Ngari, K.N., Ngunjiri, J.W., Nigatu, Y.T., Oancea, B., Oh, I., Okunga, E.W., Olusanya, J.O., Olusanya, B.O., Ong, S., Onwujekwe, O.E., Ortega-Altamirano, D.V., Ortiz, A., Soriano, J.B., Osarenotor, O., Ostojic, S.M., Vlassov, V., Pa, M., Pangaribuan, H.U., Park, E., Pasupula, D., Patel, J.R., Patel, S.K., Paternina-Caicedo, A.J., Pathak, M., Pathak, A., Patten, S.B., Tonelli, M., Weintraub, R.G., Paudel, S., Pepito, V.F., Thurston, G.D., Peprah, E.K., Pereira, J., Pescarini, J.M., Piccinelli, C., Pilgrim, T., Plass, D., Pokhrel, K.N., Prada, S.I., Prakash, V., Prakash, S., Prasad, N., Shigematsu, M., Pribadi, D.R.A., Sudaryanto, A., Rabiee, M., Radfar, A., Ranabhat, C.L., Ranta, A., Ribeiro, D.C., Rao, S., Rawaf, D.L., Rawal, L., Renjith, V., Rickard, J., Roever, L., Romoli, M., Rostamian, M., Rubagotti, E., Rwegerera, G.M., Ullah, S., Sajadi, S., Salam, N., Salomon, J.A., Salz, I., Saraswathy, S.Y., Sarmiento-Suárez, R., Sathian, B., Savic, M., Saylan, M., Schwendicke, F., Sekerija, M., Senthilkumaran, S., Sha, F., Shaheen, A.A., Shaikh, M.A., Shamsizadeh, M., Shannawaz, M., Sharma, R., Shin, J., Shiri, R., Sierpinski, R., Sigurvinsdottir, R., Silva, D.A.S., Simonetti, B., Singh, A., Sinha, D.N., Skiadaresi, E., Skou, S.T., Skryabin, V.Y., Zastrozhin, M.S., Soheili, A., Sokhan, A., Somefun, O.D., Korotkova, A.V., Soyiri, I.N., Sreeramareddy, C.T., Srinivasan, V., Sripada, K., Stovner, L.J., Stockfelt, L., Sultan, I., Swope, C.B., Vaicekonyte, R., Sykes, B.L., Szumowski, L., Tabb, K.M., Tabuchi, T., Taddele, B.W., Tahir, Z., Tareque, M.I., Tarigan, I.U., Taveira, N., Thankappan, K.R., Thomas, N., Varughese, S., Touvier, M., Tran, B.X., Tsatsakis, A., Vardavas, C., Ullah, I., Umeokonkwo, C.D., Undurraga, E.A., Upadhyay, E., Vasankari, T.J., Vasconcelos, A.N., Veisani, Y., Vidale, S., Waheed, Y., Wei, J., Wang, F., Zhao, X.G., Wang, Y., Yu, C., Zhang, Z., Wang, H., Weiss, J., Wiangkham, T., Wickramasinghe, N.D., Wu, A., Xie, Y., Xu, G., Yahyazadeh Jabbari, S., Yilgwan, C.S., Yonemoto, N., Younis, M.Z., Younker, T.P., Yu, Y., Zaidi, S., Zhang, J., Zhang, Y., Zheleva, B., Zhu, C., 2020. Global burden of 87 risk factors in 204 countries and territories, 1990–2019: a systematic analysis for the Global Burden of Disease Study 2019. The Lancet 396, 1223–1249. https://doi.org/10.1016/S0140-6736(20)30752-2

Ogen, Y., 2020. Assessing nitrogen dioxide (NO2) levels as a contributing factor to coronavirus (COVID-19) fatality. Science of the Total Environment 726, 138605. https://doi.org/10.1016/j.scitotenv.2020.138605

Peña, M., Velazquez, E., Rivera, J., Alenezi, F., Wong, C., Grigsby, M., 2017. Biomass fuel smoke exposure was associated with adverse cardiac remodeling and left ventricular dysfunction in Peru. Indoor Air 27, 737–45.

Pérez-Fargallo, A., Bienvenido-Huertas, D., Rubio-Bellido, C., Trebilcock, M., 2020. Energy poverty risk mapping methodology considering the user’s thermal adaptability: The case of Chile. Energy for Sustainable Development 58, 63–77. https://doi.org/10.1016/j.esd.2020.07.009

Pérez-Fargallo, A., Rubio-Bellido, C., Pulido-Arcas, J.A., Javier Guevara-García, Fco., 2018. Fuel Poverty Potential Risk Index in the context of climate change in Chile. Energy Policy 113, 157–170. https://doi.org/10.1016/j.enpol.2017.10.054

Pino, P., Iglesias, V., Garreaud, R., Cortés, S., Canals, M., Folch, W., Burgos, S., Levy, K., Naeher, L.P., Steenland, K., 2015. Chile confronts its environmental health future after 25 years of accelerated growth. Annals of Global Health 81, 354–367. https://doi.org/10.1016/j.aogh.2015.06.008

Pope, C.A., Turner, M.C., Burnett, R.T., Jerrett, M., Gapstur, S.M., Diver, W.R., Krewski, D., Brook, R.D., 2015. Relationships between fine particulate air pollution, cardiometabolic disorders, and cardiovascular mortality. Circulation Research 116, 108–115. https://doi.org/10.1161/CIRCRESAHA.116.305060

Pope, D., Diaz, E., Smith-Sivertsen, T., Lie, R.T., Bakke, P., Balmes, J.R., Smith, K.R., Bruce, N.G., 2015. Exposure to household air pollution from wood combustion and association with respiratory symptoms and lung function in nonsmoking women: Results from the RESPIRE trial, Guatemala. Environmental Health Perspectives 123, 285–292. https://doi.org/10.1289/ehp.1408200

Querol, X., Massagué, J., Alastuey, A., Moreno, T., Gangoiti, G., Mantilla, E., Duéguez, J.J., Escudero, M., Monfort, E., Pérez García-Pando, C., Petetin, H., Jorba, O., Vázquez, V., de la Rosa, J., Campos, A., Muñóz, M., Monge, S., Hervás, M., Javato, R., Cornide, M.J., 2021. Lessons from the COVID-19 air pollution decrease in Spain: Now what? Science of The Total Environment 779, 146380. https://doi.org/10.1016/j.scitotenv.2021.146380

R Core Team, 2017. R: A language and environment for statistical computing. R Foundation for Statistical Computing, Vienna, Austria.

Reeve, Scott, Hine, Bhullar, 2013. „This is not a burning issue for me”: How citizens justify their use of wood heaters in a city with a severe air pollution problem. Energy. Energy Policy 53, 204–11.

Reyes, R, Nelson, H., Navarro, F., Retes, C., 2015. The firewood dilemma: Human health in a broader context of well-being in Chile. Energy for Sustainable Development 28, 75–87.

Reyes, René, Nelson, H., Navarro, F., Retes, C., 2015. The firewood dilemma: Human health in a broader context of well-being in Chile. Energy for Sustainable Development 28, 75–87. https://doi.org/10.1016/j.esd.2015.07.005

Reyes, R, Schueftan, A., Ruiz, C., González, A., 2019. Controlling air pollution in a context of high energy poverty levels in southern Chile: Clean air but colder houses? Energy Policy 124, 301–311.

Reyes, René, Schueftan, A., Ruiz, C., González, A.D., 2019. Controlling air pollution in a context of high energy poverty levels in southern Chile: Clean air but colder houses? Energy Policy 124, 301–311. https://doi.org/10.1016/j.enpol.2018.10.022

Sanhueza, P.A., Torreblanca, M.A., Diaz-Robles, L.A., Schiappacasse, L.N., Silva, M.P., Astete, T.D., 2009. Particulate air pollution and health effects for cardiovascular and respiratory causes in Temuco, Chile: A wood-smoke-polluted urban area. Journal of the Air and Waste Management Association 59, 1481–1488. https://doi.org/10.3155/1047-3289.59.12.1481

Schueftan, A., González, A., 2013. Reduction of firewood consumption by households in south-central Chile associated with energy efficiency programs. Energy Policy 63, 823–832.

Schueftan, A., González, A.D., 2015. Proposals to enhance thermal efficiency programs and air pollution control in south-central Chile. Energy Policy 79, 48–57. https://doi.org/10.1016/j.enpol.2015.01.008.

Schueftan, A., Sommerhoff, J., González, A., 2016. Firewood demand and energy policy in south-central Chile. Energy for Sustainable Development 33, 26–35. https://doi.org/10.1016/j.esd.2016.04.004.

Setti, L., Passarini, F., Gennaro, G. De, Al, E., 2020. Searching for SARS-COV-2 on Particulate Matter: A Possible Early Indicator of COVID-19 Epidemic Recurrence. International Journal of Environmental Research and Public Health 17.

Shupler, M., Hystad, P., Birch, A., Miller-Lionberg, D., Jeronimo, M., Arku, R.E., Chu, Y.L., Mushtaha, M., Heenan, L., Rangarajan, S., Seron, P., Lanas, F., Cazor, F., Lopez-Jaramillo, P., Camacho, P.A., Perez, M., Yeates, K., West, N., Ncube, T., Ncube, B., Chifamba, J., Yusuf, R., Khan, A., Hu, B., Liu, X., Wei, L., Tse, L.A., Mohan, D., Kumar, P., Gupta, R., Mohan, I., Jayachitra, K.G., Mony, P.K., Rammohan, K., Nair, S., Lakshmi, P.V.M., Sagar, V., Khawaja, R., Iqbal, R., Kazmi, K., Yusuf, S., Brauer, M., 2020a. Household and personal air pollution exposure measurements from 120 communities in eight countries: results from the PURE-AIR study. The Lancet Planetary Health 4, e451–e462. https://doi.org/10.1016/S2542-5196(20)30197-2

Shupler, M., Mwitari, J., Gohole, A., de Cuevas, R.A., Puzzolo, E., Čukić, I., Nix, E., Pope, D., 2020b. COVID- 19 Lockdown in a Kenyan informal settlement: Impacts on household energy and food security. medRxiv. https://doi.org/10.1101/2020.05.27.20115113

Siddharthan, T., Grigsby, M.R., Goodman, D., Chowdhury, M., Rubinstein, A., Irazola, V., Gutierrez, L., Jaime Miranda, J., Bernabe-Ortiz, A., Alam, D., Kirenga, B., Jones, R., Van Gemert, F., Wise, R.A., Checkley, W., 2018. Association between household air pollution exposure and chronic obstructive pulmonary disease outcomes in 13 low- and middle-income country settings. American Journal of Respiratory and Critical Care Medicine 197, 611–620. https://doi.org/10.1164/rccm.201709-1861OC

SINCA, 2015. Sistema de Información Nacional de Calidad del Aire, Ministerio del Medio Ambiente, Gobierno de Chile.

Snyder, E.G., Watkins, T.H., Solomon, P.A., Thoma, E.D., Williams, R.W., Hagler, G.S.W., Shelow, D., Hindin, D.A., Kilaru, V.J., Preuss, P.W., 2013. The changing paradigm of air pollution monitoring. Environmental Science and Technology 47, 11369–11377. https://doi.org/10.1021/es4022602

State of Global Air, 2020. Health Effects Institute [WWW Document]. URL https://www.stateofglobalair.org/

Tagle, M., Rojas, F., Reyes, F., Vásquez, Y., Hallgren, F., Lindén, J., Kolev, D., Watne, Å. K., Oyola, P., 2020. Field performance of a low-cost sensor in the monitoring of particulate matter in Santiago, Chile. Environ Monit Assess 192, 171. https://doi.org/10.1007/s10661-020-8118-4

Tariq, A., Undurraga, E.A., Laborde, C.C., Vogt-Geisse, K., Luo, R., Rothenberg, R., Chowell, G., 2021. Transmission dynamics and control of covid-19 in chile, march-october, 2020. PLoS Neglected Tropical Diseases 15, 1–20. https://doi.org/10.1371/journal.pntd.0009070

Thu, T.P.B., Ngoc, P.N.H., Hai, N.M., Tuan, L.A., 2020. Effect of the social distancing measures on the spread of COVID-19 in 10 highly infected countries. Science of the Total Environment 742, 140430. https://doi.org/10.1016/j.scitotenv.2020.140430

Tsapakis, M., Evaggelia Lagoudaki, Stephanou, E.G., Kavouras, I.G., Koutrakis, P., Oyola, P., Baer, D. von, 2002. The composition and sources of PM2. 5 organic aerosol in two urban areas of Chile. Atmospheric Environment 36, 3851–3863.

Umaña-Hermosilla, B., de la Fuente-Mella, H., Elórtegui-Gómez, C., Fonseca-Fuentes, M., 2020. Multinomial logistic regression to estimate and predict the perceptions of individuals and companies in the face of the covid-19 pandemic in the Ñuble region, Chile. Sustainability (Switzerland) 12, 1–20. https://doi.org/10.3390/su12229553

Undurraga, E.A., Chowell, G., Mizumoto, K., 2021. COVID-19 case fatality risk by age and gender in a high testing setting in Latin America: Chile, March–August 2020. Infectious Diseases of Poverty 10, 1–11. https://doi.org/10.1186/s40249-020-00785-1

Vergara, L., 2019. Medianización social y transformaciones residenciales recientes en ciudades de La Araucanía. Cultura-hombre-sociedad 29, 36–60. https://doi.org/10.7770/0719-2789.2019.cuhso.04a03

Wu, X., Nethery, R.C., Sabath, B.M., Al, E., 2020. Exposure to Air Pollution and COVID-19 Mortality in the United States: A Nationwide Cross-Sectional Study. MedRxiv Preprint.

Zhu, Y., Xie, J., Huang, F., Cao, L., 2020. Association between short-term exposure to air pollution and COVID-19 infection: Evidence from China. Science of the Total Environment 727, 138704. https://doi.org/10.1016/j.scitotenv.2020.138704

