## Supplemental Information for "Energy Insecurity Influences Urban Outdoor Air Pollution Levels during COVID-19 Lockdown in South-Central Chile"

#### Seasonal fluctuations in fine particulate matter concentrations

Concentrations of fine particulate matter ($PM_{2.5}$) are significantly higher in Temuco during the colder months (April- August) of the year compared with warmer months (January-March) when residential wood heating is not typically needed. While there was a moderate correlation between heating degree days and average 24-hour $PM_{2.5}$ concentrations (Figure 2 in main text), $PM_{2.5}$ concentrations in April 2020 and September 2020 were similar despite September having colder temperatures (higher heating degree days) (Figure S1). As April 2020 coincided with the strictest quarantine measures during lockdown in Temuco, while September 2020 fell under an 'initial opening' phase, it is likely that household confinement in April 2020 was a driver of $PM_{2.5}$ concentrations increasing above typical levels in many areas of Temuco. This finding is supported by a significantly higher fine fraction of $PM_{10}$ registered to many monitoring stations in winter months of 2020 compared with 2019 (Figure 9 in main text).

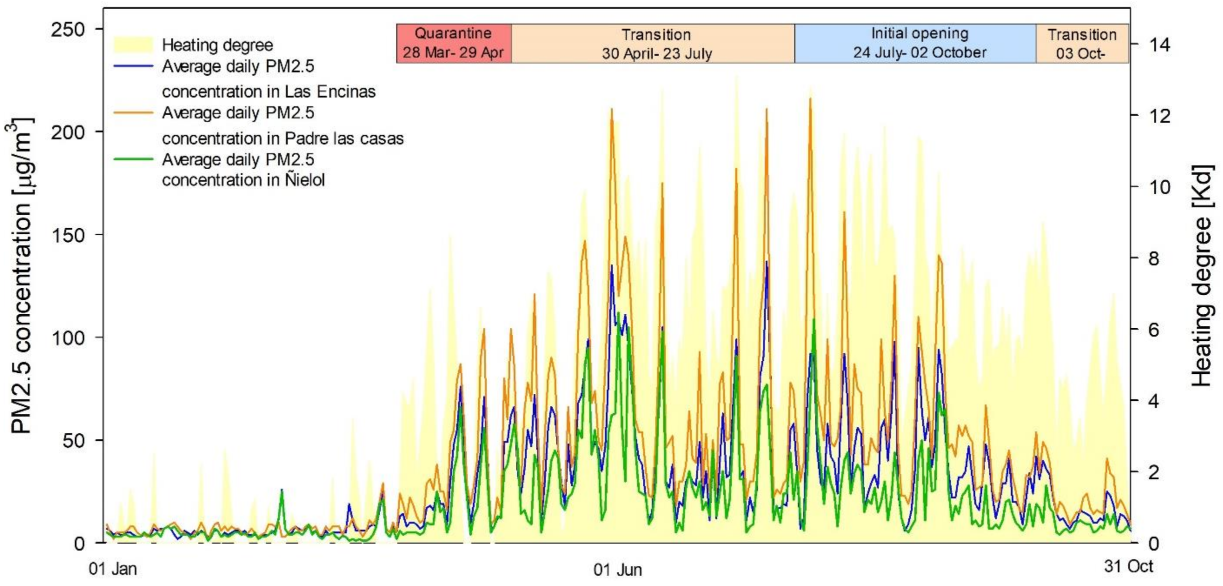


**Figure S1**. Average 24-hour PM2.5 concentration and heating degrees for January-October 2020

In addition to large seasonal variation, $PM_{2.5}$ concentrations vary substantially over the course of the day. An increase in $PM_{2.5}$ concentrations typically occurs at night, between 5:00 PM–9:00 AM, when individuals are home from work and burning wood to heat their homes (Figure S2).

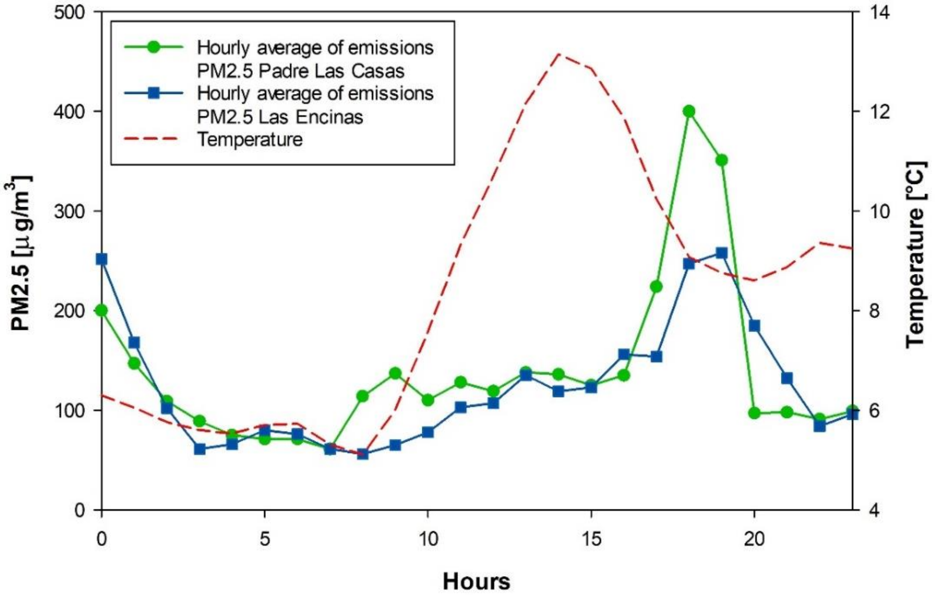


**Figure S2**. Example of average hourly $PM_{2.5}$ concentrations and temperatures at Las Encinas and Padre Las Casas (May 25, 2019)

### Calibration of Sensirion SPS30 sensors

The coefficient of determination between the Sensirion SPS30 sensors deployed at Pedro Valdivia, Pueblo Nuevo and Centro and the Grimm laser aerosol spectrometer 11-E during an 8-hour calibration was 0.92 for $PM_{2.5}$ and 0.96 for $PM_{10}$ (Figures S3 and S4, respectively). The sensors performed best at concentrations below 500 µg/m$^3$, corresponding to all measurements in the study.

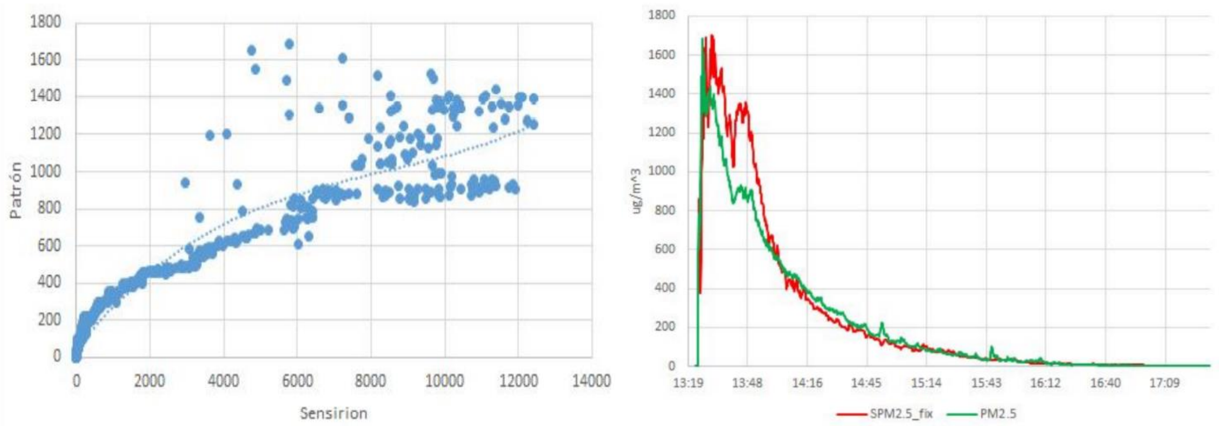


**Figure S3.** (left) Calibration curve for $PM_{2.5}$ concentrations between Sensirion SPS30 sensors (x-axis) and Grimm laser aerosol spectrometer 11-E (y-axis) (right) Corrected Sensirion SPS30 sensor $PM_{2.5}$ measurements (SPM2.5_fix) alongside Grimm laser aerosol spectrometer 11-E $PM_{2.5}$ concentrations (PM2.5)

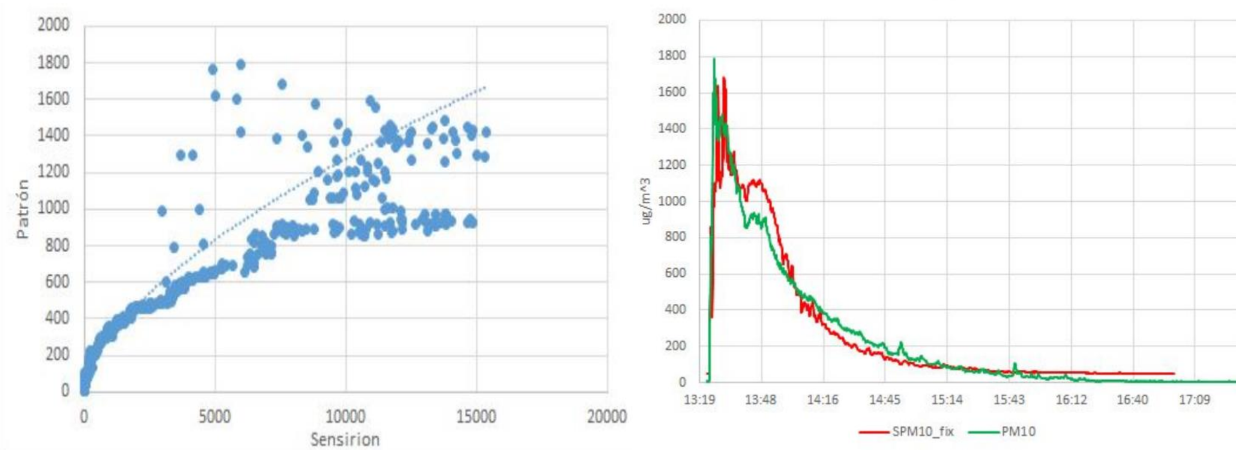


**Figure S4.** (left) Calibration curve for $PM_{10}$ concentrations between Sensirion SPS30 sensors (x-axis) and Grimm laser aerosol spectrometer 11-E (y-axis) (right) Corrected Sensirion SPS30 sensor $PM_{10}$ measurements (PM10_fix) alongside Grimm laser aerosol spectrometer 11-E $PM_{10}$ concentrations (PM10)

### Critical episodes

'Critical episodes' occur when the 24-hour moving average ambient $PM_{2.5}$ concentration is greater than 79 µg/m$^3$ or the 24-hour moving average ambient $PM_{10}$ concentration is greater than 195 µg/m$^3$. Critical episodes are used as a tool to enforce temporary bans on residential wood heating, and notify the public about the dangers of outdoor activity on days with poor ambient air quality. Critical episodes are categorized into 'alert', 'pre-emergency' or 'emergency' (Table S1). 'Non-critical' episodes are characterized as 'good' if 24-hour moving average $PM_{2.5}$ concentration in compliance the WHO annual target level (25 µg/m$^3$), or 'regular' if 24-hour moving average $PM_{2.5}$ concentration is between 25-79 µg/m$^3$.

**Table S1**. Categorization of air quality according to concentration levels of particulate matter

| Air quality | $PM_{10}$ (µg/m$^3$) | $PM_{2.5}$ (µg/m$^3$) | Category |
|---|---|---|---|
| Good | 0-149 | 0-25 | Non-critical episodes |
| Regular | 150-194 | 26-79 | |
| Alert | 195-239 | 80-109 | Critical episodes |
| Pre-emergency | 240-329 | 110-169 | |
| Emergency | ≥330 | ≥170 | |

From March 1st to September 30th in 2019 and 2020, all critical episodes were initiated by elevated $PM_{2.5}$ levels; $PM_{10}$ concentrations did not exceed the threshold needed to trigger a critical episode in either year. The total number of critical episodes in residential areas (Las Encinas and Padre Las Casas) in 2019 and 2020 were twice and four times greater, respectively, than in the commercial area of Ñielol (Table S2).

During COVID-19 lockdown in 2020, the number of ‘alert’ critical episodes detected by all beta attenuation monitors increased substantially ((Ñielol (40%), Padre Las Casas (56%) and Las Encinas (107%)), with the largest increase occurring in the middle-income residential neighborhood of Las Encinas. The total number of critical episodes increased by 46% in Las Encinas, 38% in Ñielol and 16% in Padre Las Casas.

**Table S2**. Number of critical episodes in 2019 and 2020 based on beta attenuation monitor measurements

| | Ñielol | | Las Encinas | | Padre Las Casas | |
|---|---|---|---|---|---|---|
| | 2019 | 2020 | 2019 | 2020 | 2019 | 2020 |
| Alert | 10 | 14 | 14 | 29 | 23 | 36 |
| Pre-emergency | 3 | 3 | 12 | 8 | 29 | 24 |
| Emergency | 0 | 0 | 1 | 1 | 14 | 14 |
| Total critical episodes | 13 | 17 | 27 | 38 | 66 | 74 |
